# Linking transmission dynamics and economic evaluation to assess schistosomiasis programme strategies across districts in Malawi

**DOI:** 10.64898/2026.05.06.26352519

**Authors:** Tara Danielle Mangal, Tim Colbourn, Andrew Phillips, Joseph Mfutso-Bengo, Pemphero Mphamba, Sakshi Mohan, Rachel Murray-Watson, Dominic Nkhoma, Eva Janoušková, Bingling She, Paul Revill, Timothy Hallett

**Author notes:** Corresponding author: Dr T. Mangal. authors listed alphabetically (Tim Colbourn, Andrew Phillips, Joseph Mfutso-Bengo, Pemphero Mphamba, Sakshi Mohan, Rachel Murray-Watson, Dominic Nkhoma, Eva Janoušková, Bingling She).

## Abstract

**Background:** Preventive chemotherapy targeting school-aged children has substantially reduced schistosomiasis morbidity, however, a key strategic tension remains between sustaining morbidity control and pursuing transmission elimination, particularly in settings characterised by heterogeneous transmission dynamics and persistent adult infection reservoirs. We developed a health system-integrated transmission and economic evaluation framework to identify optimal age-targeting and district-level prioritisation, providing a basis for determining when elimination-focused approaches offer advantages over morbidity reduction alone.

**Methods:** The *Thanzi la Onse* individual-based model was used to evaluate alternative age-targeted mass drug administration (MDA) strategies for *Schistosoma haematobium* and *Schistosoma mansoni* across all 32 districts of Malawi from 2024-2050. Strategies included treatment of school-aged children (MDA-SAC), pre-school and school-aged children (MDA-PSAC+SAC), and community-wide treatment (all ages). Health outcomes included person-years with any infection (PY), disability-adjusted life years (DALYs), probability of elimination (defined as reaching <2% prevalence of infection in all ages). The cost-effectiveness was evaluated using incremental cost-effectiveness ratios (ICERs), net health benefit (NHB), and by quantifying the maximum costs available for implementation, using a cost-effectiveness threshld for Malawi of 88 USD per DALY averted.

**Findings:** In the absence of MDA, the majority of the infection burden over 2024–2050 would be concentrated in adults aged 15 years and older (219.6 million person-years [PY], 95% CI 215.4–223.4), compared with 72.8 million PY (95% CI 71.5–74.3) among school-aged children (SAC) and 25.5 million PY (95% CI 25.1– 26.2) among preschool-aged children. Annual MDA-SAC would avert approximately 18.0 million DALYs (95% CI 17.6–18.4) between 2025 and 2050 and would be highly cost-effective nationally (ICER 4.76 USD/DALY, 95% CI 4.47–4.95). Across districts, ICERs were highly variable; 25 of 32 districts were cost-effective in ≥90% of runs and 29 of 32 in ≥50% of runs.

Expanding treatment to include preschool-aged children (MDA PSAC+SAC) would produce modest additional gains (additional 44,500 DALYs averted) but with substantially higher costs (national ICER 606 USD/DALY, 95% CI 472–695), being dominated in 22 districts and cost-effective only in the high-burden Likoma district. Community-wide MDA would achieve elimination for both species in all districts by 2030 and avert a further 98,000 DALYs; nationally it would be cost-saving relative to PSAC+SAC although outcomes were heterogeneous, with this strategy being cost-saving in 11 high-prevalence districts (2023 prevalence range 13.7 – 41.5%) but dominated (in >80% of model runs) in 16 others. Threshold analyses of maximum implementation costs indicated substantial cost margins in high-burden districts, with cost-effectiveness maintained up to approximately 25–38 USD per treatment.

**Interpretation:** The choice of schistosomiasis strategies should depend on whether programmes prioritise short-term morbidity reduction or long-term elimination, as well as the local disease burden and the prevailing cost of service delivery. Integrating district-level transmission dynamics with opportunity-cost–based economic evaluation reveals when broader coverage is justified and provides a framework for designing fiscally grounded elimination pathways in heterogeneous endemic settings.

## Introduction

Schistosomiasis remains a persistent public health challenge across sub-Saharan Africa, contributing to chronic morbidity, impaired child development, and reduced economic productivity.(Hotez and Kamath 2009, World Health Organization 2022) Despite long-standing mass drug administration (MDA) campaigns with praziquantel, progress towards sustained control and elimination has been uneven. National programmes often rely on uniform treatment schedules that do not fully reflect spatial heterogeneity in transmission intensity, age-specific exposure, or health-system capacity.(World Health Organization 2006, World Health Organization 2011) As a result, substantial reservoirs of infection persist in high-burden districts and reinfection remains common, particularly where environmental and behavioural risk factors are entrenched.(Rollinson, Knopp et al. 2013, Colley, Bustinduy et al. 2014)

Mechanistic transmission models have been instrumental in improving understanding of schistosomiasis epidemiology, including the effects of repeated MDA, age-dependent exposure, and the conditions required for sustained reductions in transmission. Dynamic, age-structured deterministic models have been used to explore reinfection dynamics and intervention thresholds (Lo, Lai et al. 2016, Gurarie, Lo et al. 2018) while geostatistical analyses have highlighted pronounced district-level heterogeneity and the persistence of local hotspots even where national averages appear modest.(Schur, Vounatsou et al. 2012, Lai, Biedermann et al. 2015) More recently, individual-based models have expanded the modelling toolkit by simulating complex host-level processes across a range of epidemiological settings.(Graham, Ayabina et al. 2021) Water, sanitation and hygiene (WASH) interventions are increasingly recognised as structural complements to preventive chemotherapy because they reduce environmental exposure and reinfection risk.(1) Evidence syntheses and modelling studies suggest that WASH can enhance long-term transmission reduction and support elimination alongside MDA, although effects are gradual and context-dependent, with chemotherapy remaining central for short-term morbidity control.(2–5)

However, moving from morbidity control to elimination requires analytical frameworks that link biological transmission processes with the health-system pathways through which diagnosis, treatment, and commodity availability shape real-world programme performance. Economic evaluations and modelling studies have provided strong evidence supporting preventive chemotherapy and alternative delivery strategies, including integrated and community-wide approaches, and systematic reviews indicate that schistosomiasis control is generally cost-effective across diverse settings.(6–8) In settings where *Schistosoma mansoni* and *Schistosoma haematobium* circulate concurrently, there remains scope for integrated transmission-economic analyses that evaluate alternative age-targeting strategies across heterogeneous geographies to inform context-specific pathways from control towards elimination.

Malawi remains co-endemic for *S. haematobium* and *S. mansoni*, with marked district heterogeneity in transmission and ongoing burden despite repeated preventive chemotherapy campaigns, making it a relevant setting in which to evaluate geographically targeted elimination strategies.(9) To address this, we developed a health-system–integrated schistosomiasis transmission and economic evaluation framework within the *Thanzi La Onse* (TLO) platform.(Hallett, Mangal et al. 2024) This framework builds on prior mechanistic and individual-based models by incorporating age-structured and species-specific transmission, care-seeking and diagnostic pathways, WASH transitions, district-level heterogeneity, and linking epidemiological outputs to economic decision metrics including incremental cost-effectiveness ratios (ICERs), net health benefit (NHB), and maximum allowable costs (MAC).

We assessed the district-level health impact, cost-effectiveness, and elimination feasibility of alternative age-targeted MDA strategies, treating school-aged children (SAC), preschool- and school-aged children (PSAC+SAC), or all individuals aged ≥2 years (MDA All) under varying WASH trajectories in Malawi. Our objectives were: (i) to quantify district-specific health gains; (ii) to examine the likelihood of achieving elimination thresholds for *S. haematobium* and *S. mansoni* by 2050, and (iii) estimate the economic value and fiscal feasibility of each strategy.

## Methods

### Study design and modelling framework

The Thanzi la Onse (TLO) individual-based model simulates population dynamics, infection processes, care-seeking behaviour, diagnostic and treatment pathways, and commodity availability within each of Malawi’s 32 administrative districts.(Hallett, Phillips et al. 2024) The framework was extended to incorporate species-specific transmission of *S. haematobium* and *S. mansoni*, WASH-driven exposure, and district-level heterogeneity in baseline prevalence and environmental risk. TLO represents real-world health-system processes through explicitly modelled care pathways. Individuals who seek healthcare can encounter probabilistic delays and constraints arising from staffing, diagnostics, supply chains, and facility-level commodity availability. These influence both the timing and likelihood of treatment delivery and are parameterised using structured facility surveys, national administrative data, and empirical care-seeking patterns. Further details on all methods, interactions, parameter values, and data sources are provided in the appendix.

### Schistosomiasis transmission model

For each species (*d*) and district (*i*), transmission intensity was determined using a fixed transmission coefficient (*κ*), encapsulating ecological and behavioural determinants of exposure, including snail population dynamics, environmental survival of free-living stages and human-water contact. Transmission was simulated independently within administrative districts, with no assumed movement between districts. At monthly time-steps, individual worm burdens were aggregated to form a district-level reservoir of infection representing environmental contamination. For district *i* and age-group *j*, the mean worm burden 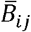 was calculated across all individuals, and the total reservoir defined as:

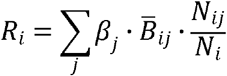

where *β*_*j*_ denotes age-specific exposure and *N*_*i,j*_ the corresponding population size. A small prevalence-dependent adjustment was applied to this reservoir to avoid unrealistically abrupt decline following intensive intervention while preserving the potential for elimination at low transmission. The district force of infection was then defined as:

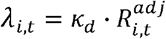

Baseline susceptibility (*s*_*k*_) in each district was calibrated to reproduce observed prevalence among school-aged children and preferentially assigned to individuals without improved sanitation. Each individual was assigned a worm-harbouring propensity (*h*_*k*_) drawn from a gamma distribution to reproduce aggregation of infection. The effective acquisition rate for individual k was defined as:

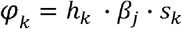

New worm acquisition followed a Poisson process proportional to the district force of infection and the individual acquisition rate. Establishment of new worms was density-dependent, limited by existing host worm burden and species-specific fecundity, with a stochastic maturation delay of 30-55 days before worms began producing eggs.

Simulations were initialised by distributing worms across individuals in proportion to their exposure risk and harbouring propensities to reproduce district- and age-specific mean worm burdens. Infection intensity was derived from the simulated worm burdens using the World Health Organization egg-count thresholds into equivalent worm numbers for each species, classifying infections into light, moderate and heavy infection. Health loss was quantified as years lived with disability by assigning intensity-specific disability weights multiplied by the duration spent with infection.

### Health system interactions, treatment and WASH effects

Clinical care-seeking was modelled as a probabilistic process triggered by infection severity for individuals with moderate or heavy worm burdens. Diagnostic assessment followed national clinical guidelines, with urine filtration used for urinary presentations and Kato-Katz stool microscopy otherwise. Test sensitivity was intensity-dependent, with low sensitivity of light infections. Individuals with a positive diagnosis were eligible for praziquantel treatment, subject to facility-level constraints on drug stock and healthcare worker time. Praziquantel was administered at age-appropriate doses, with species-specific adult worm clearance probabilities applied to reduce individual worm burdens. MDA campaigns operated independently of routine care without diagnostic screening, and applied the same probabilistic treatment efficacy to eligible age-groups.

WASH status was represented at the individual level and initialised using nationally representative survey distributions stratified by residence and household wealth.(10) Continued improvements in sanitation and access to clean water followed historical trends and reduced the probability of remaining susceptible to infection through a composite reduction of 40% in exposure risk applied uniformly across age-groups and species.(3)

### Intervention scenarios

We evaluated five intervention scenarios against the baseline assuming no MDA: (1) MDA targeting school-aged children (SAC; 5-14 years); (2) MDA expanded to include preschool-aged and school-aged children (PSAC+SAC; 2-14 years); (3) community-wide MDA for all individuals ≥ 2 years; (4) WASH scale-up only. Coverage was assumed to be 80% across eligible age-groups.

Sensitivity analyses examined alternatives to key assumptions:

i. alternative WASH trajectories, including “Pause WASH” where WASH coverage was held constant at 2023 levels, and “Scale-up WASH”, representing an immediate improvement across all WASH attributes
ii. assigning non-zero disability weights to light infections, which are otherwise assumed to carry no symptomatic burden in the base case.

### Epidemiological outcomes

We estimated person-years (PY) of infection by species, age group, and district; DALYs based on species-specific disability contributions and intensity categories; prevalence at each intensity; and time to elimination, defined as <2% prevalence for both species – a threshold identified through model testing below which stochastic rebound was unlikely to occur. Where prevalence fell below the elimination threshold for two consecutive years, the model ceased further MDA in subsequent years. Disability weights, intensity thresholds, and sensitivity analyses are described in the appendix.

### Economic evaluation

Incremental cost-effectiveness ratios (ICERs), defined as the ratio of the difference in costs to the difference in health outcomes between competing strategies (i.e., cost per disability-adjusted life year (DALY) averted), were calculated using consumables costs alongside the direct financial costs for implementation (including programme management, community sensitisation, training and drug delivery), reflecting the variable component of expenditure that scales directly with the number of individuals treated. (Morozoff, Avokpaho et al. 2022) Consumables comprised praziquantel tablets costed using nationally sourced unit prices, with required quantities determined by weight-based dosing derived from age-specific body weight distributions.(11) As the implementation costs were derived using empirical estimates from small-scale operational studies and may not scale linearly with coverage, an additional scenario assuming 50% implementation costs alongside consumables was included.

To characterise fiscal feasibility under each strategy, we report the maximum allowable delivery cost (MADC), defined as the additional implementation cost (beyond consumables) that can be incurred while remaining cost-effective at Malawi’s opportunity-cost threshold. ICERs and net health benefit (NHB), defined as the monetised health gain (in DALYs averted) minus incremental costs scaled by the cost-effectiveness threshold, were calculated using Malawi’s opportunity-cost–based threshold inflated to 2025 (CET 88 USD per DALY averted).(12) NHB integrates health gains and incremental costs to determine the preferred strategy in each district using no MDA as the comparator. Full costing tables, discounting assumptions, and sensitivity analyses are presented in the appendix.

### Calibration, data sources, and uncertainty

Species-specific endemic equilibria were first estimated in a fully susceptible population to characterise the mapping between susceptibility and prevalence. District-level prevalence of *S. haematobium* and *S. mansoni* was then calibrated to pre-MDA survey data collected between 1982 and 2015, and combined with the worm aggregation distribution to estimate, for each district, the proportion susceptible and mean worm burden used to initialise simulations.(World Health Organization Regional Office for Africa 2024) Model validation was performed independently using compiled district and site-level survey data from 2019. Simulated district prevalence for both species lay within or close to observed uncertainty ranges in most locations, with remaining discrepancies small and consistent with expected sampling variability between site-level observations and district-level model outputs.

### Population and Time Horizon

A representative open cohort of 3,000 individuals per district (96,000 total) was simulated from 2010 to 2050. Historical MDA campaigns were represented according to available data up to 2022. From 1^st^ January 2023, alternative intervention strategies were implemented, with MDA programmes occurring at the mid-point of each year. Each scenario was replicated 10 times to account for stochastic variability. All outputs are presented as mean estimates with 95% confidence intervals (CI), aggregated nationally (weighted by district population) or at district-level. All analyses were conducted in Python v3.11.11. The code used for analysis and figure generation is publicly available on Github at https://github.com/UCL/TLOmodel.git under the tag “schisto-analysis-v1.0”.

### Ethical approval

The Thanzi La Onse project received ethical approval from the College of Medicine Malawi Research Ethics Committee (COMREC, P.10/19/2820) for the use of publicly accessible and anonymised secondary data. No data were used requiring individual informed consent.

## Results

### District-specific health gains under alternative MDA strategies

Substantial epidemiological heterogeneity was observed at baseline (2023) for both *S. haematobium* and *S. mansoni*, with end-of-year prevalence ranging from <1% in several low-burden districts to nearly 40%. Adults represented the largest infection reservoir in 2024, contributing 3.20 million (95% CI: 3.04–3.23) PY compared with 1.11 million (95% CI: 1.08–1.15) PY in SAC and 0.52 million (95% CI: 0.49–0.54) PY in preschool-aged children (PSAC) (Figure 1, Appendix Table 5). Heavy-intensity infections were less frequent but geographically concentrated, with five districts jointly contributing 31% and 27% of the national heavy-intensity burden in both adults and children. WASH conditions were heterogeneous with a lack of functional handwashing facilities, affecting > 60% of households in most districts, while lack of access to clean drinking water affected between 8–12% of households (Appendix Figure 1).

**Figure 1.**
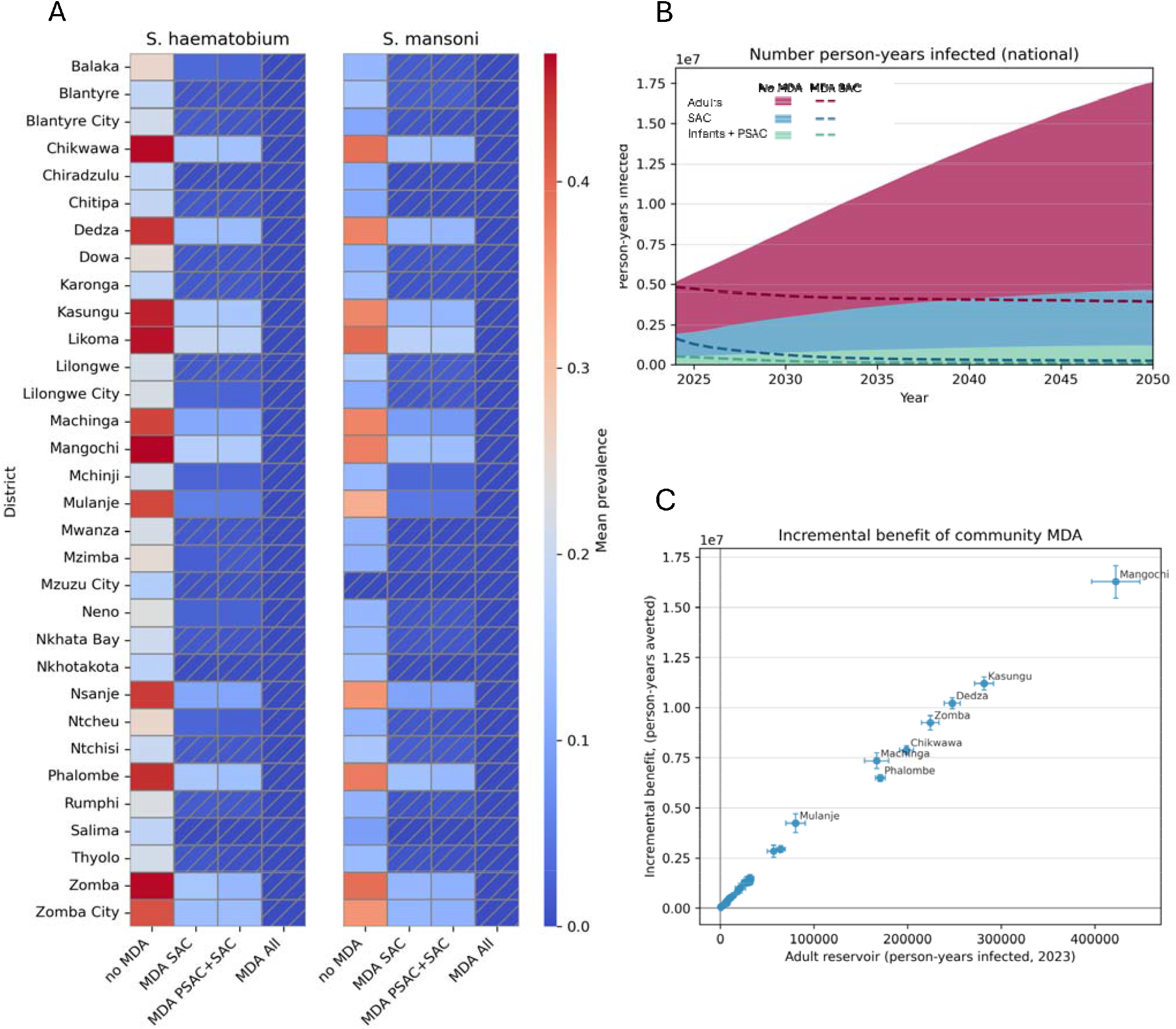
(A) The estimated mean prevalence of *S. haematobium* and *S. mansoni* in 2050 under each MDA strategy. Hatched cells highlight those districts which would achieve elimination (<2% prevalence of both species). (B) The number of person-years infected with either species by age-group. The shaded areas show projections with no MDA, dashed lines show MDA for school-aged children once annually. (C) The incremental benefit of expanding MDA to adults. Each point represents one district with the reservoir of infection in adults in 2023 (baseline) plotted against the incremental benefit (number of person-years of infection averted in adults over 2024-2050). Error bars show the 95% confidence intervals.

Over 2024-2050, the cumulative infection burden without MDA was 219.6 million PY (95% CI: 215.4–223.4) in adults, 72.8 million PY (95% CI: 71.5–74.3) in SAC and 25.5 million PY (95% CI: 25.1–26.2) in infants plus PSAC. If WASH scale-up was halted (Pause WASH), the adult burden was sustained at 219.2 million PY (95% CI: 214.2–223.4), with minimal increases also evident in younger groups. Under Scale-up WASH, infection burden declined modestly but remained substantial (adults 215.4 million PY, 95% CI: 209.5–219.8; infants plus PSAC 23.0 million PY, 95% CI: 22.3–23.8; SAC 68.1 million PY, 95% CI: 66.2–69.9).

All MDA strategies reduced infection, with effects increasing with programme scope (Figure 1A). With MDA SAC, trajectories showed sustained reductions in infection across age groups relative to no MDA (Figure 1B), averting 62.2 million SAC PY (88.0%, 95% CI: 87.7–88.4%) and 115.9 million adult PY (54.6%, 95% CI: 53.5– 55.8%) through indirect reductions in environmental contamination, although adult infection persisted in most districts. MDA PSAC+SAC reduced 90.7% (95% CI: 90.4–91.1) of SAC PY and 91.0% (95% CI: 90.6–91.4) of infant plus PSAC PY, but had a smaller effect on adults (56.0%, 95% CI: 55.0–57.2), MDA All produced the largest reductions across all ages, averting >208 million adult PY, 67.8 million SAC PY and 23.4 million PSAC PY (96-98% reductions).

These reductions translated into large improvements in population health. MDA SAC averted approximately 18.0 million DALYs between 2025 and 2050 (95% CI 17.6–18.4). Extending treatment to PSAC averted an additional 44,500 DALYs, while expanding to adults yielded a further 98,000 DALYs beyond PSAC+SAC. In this base case, the majority of infection were light intensity, contributing no DALYs. Assigning a small disability weight (0.006) to light infections increased absolute burden estimates but did not alter the relative ranking of strategies, with 18.55 million, 18.64 million and 19.22 million DALYs averted through MDA SAC, PSAC+SAC and MDA All respectively. Absolute DALYs averted through MDA were lower under the most ambitious WASH scale-up (MDA All: 16.15 million [95% CI 15.71–16.59] vs 18.14 million [17.76–18.52] under continued WASH), but the relative ranking of strategies and incremental differences between targeting approaches remained unchanged (see Appendix Table 9).

The incremental benefit of expanding to adults was strongly and approximately linearly associated with the baseline adult reservoir of infection (Figure 1C). Districts with larger adult reservoirs in 2023 achieved larger incremental reductions in PY infected over 2024-2050 when moving from paediatric-only strategies to community-wide MDA.

### Feasibility and heterogeneity of achieving elimination thresholds

All strategies eliminated heavy-intensity infections nationally by 2030, however, elimination (defined as overall prevalence <2%), varied by strategy, species, and district (Figure 1A, Figure 2). MDA All was the only approach achieving elimination in all 32 districts for both *S. haematobium* and *S. mansoni* by 2030. In contrast, paediatric-focused strategies produced slower and more heterogeneous declines. Under MDA SAC, fewer than half of districts reached elimination for *S. haematobium* by 2050. Performance was somewhat better for *S. mansoni*, but many districts remained above the threshold by 2050. Districts with the highest baseline burden showed persistent residual transmission in the absence of adult treatment. These patterns were driven primarily by the large adult reservoir of infection; even under high paediatric coverage, adult PY fell by less than 60%, allowing sustained transmission.

**Figure 2.**
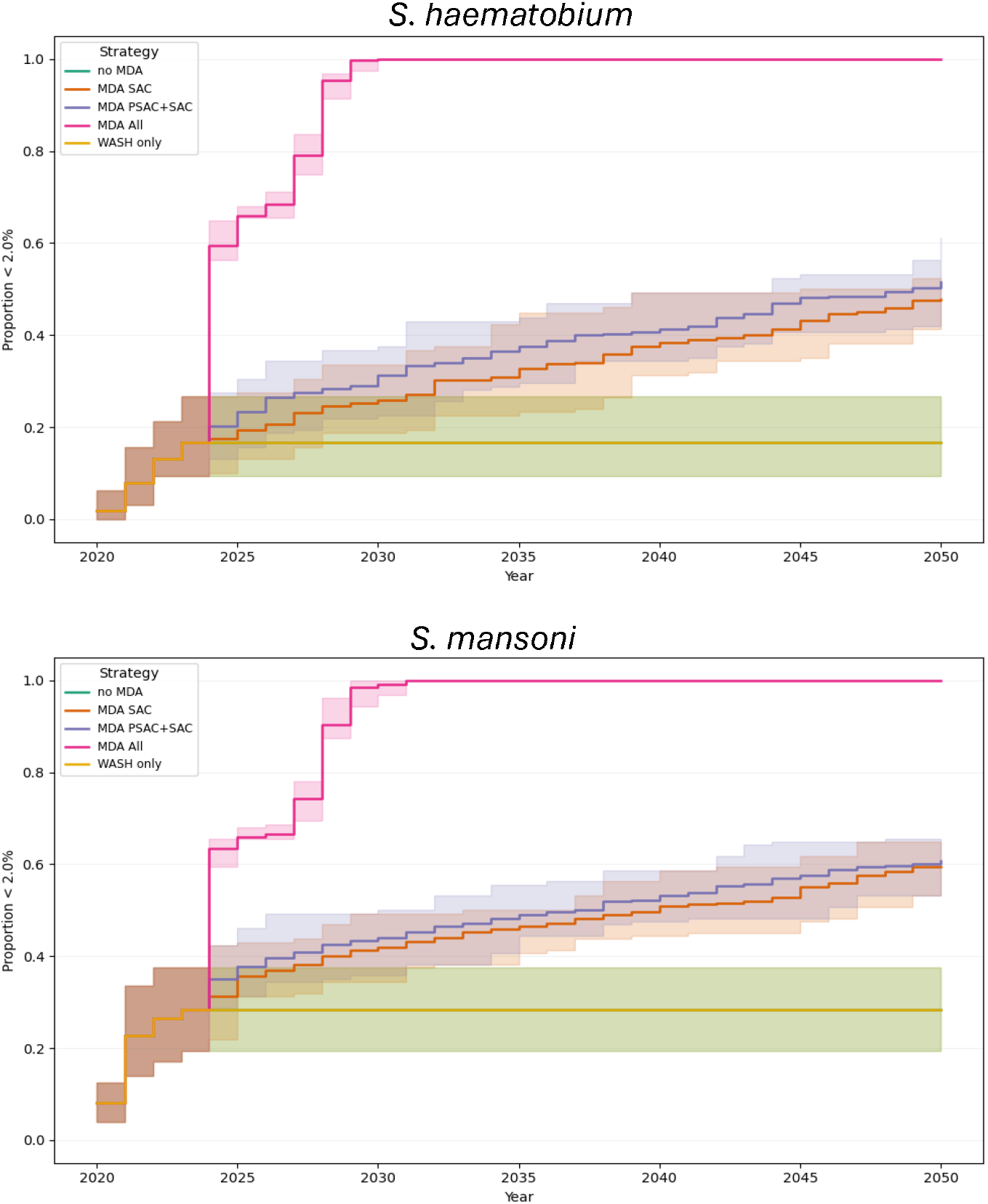
Proportion of districts achieving the <2% prevalence threshold under alternative MDA strategies for *S. haematobium* (upper figure) and *S. mansoni* (lower figure). *Lines show the projected proportion of districts falling below the 2% threshold each year from 2020–2050. Shaded regions denote confidence intervals due to stochasticity. Results are presented for no MDA, MDA SAC, MDA PSAC+SAC, MDA All and WASH-only counterfactuals*.

WASH trajectories influenced absolute prevalence but did not materially alter elimination feasibility. Scale-up accelerated declines marginally, enabling one additional district (Neno) to reach elimination for each species under MDA PSAC+SAC by 2050 relative to Continue WASH, while the Pause WASH scenario had negligible impact.

### Cost-effectiveness, net health benefit and maximum allowable costs

Over 2024–2050, total delivery costs were 166.73 million USD (95% CI 165.18–168.90 million) for MDA SAC, 220.91 million USD (95% CI 219.02–223.99 million) for MDA PSAC+SAC and 702.24 million USD (699.95– 707.81 million) for MDA All (Appendix Table 6).

Compared with no MDA, MDA SAC was highly cost-effective (ICER 4.76 USD/DALY, 95% CI 4.47-4.95) and was below the 88 USD/DALY CET in ≥90% of runs in 25 of 32 districts (Figure 3); in 29 districts it was cost-effective in at least 50% of runs. The mean district NHB for MDA SAC relative to no MDA was positive in 31/32 districts, with Mzuzu City the only district with negative mean NHB (Figure 4A), although 7 districts had uncertainty intervals crossing zero indicating uncertainty around cost-effectiveness.

**Figure 3.**
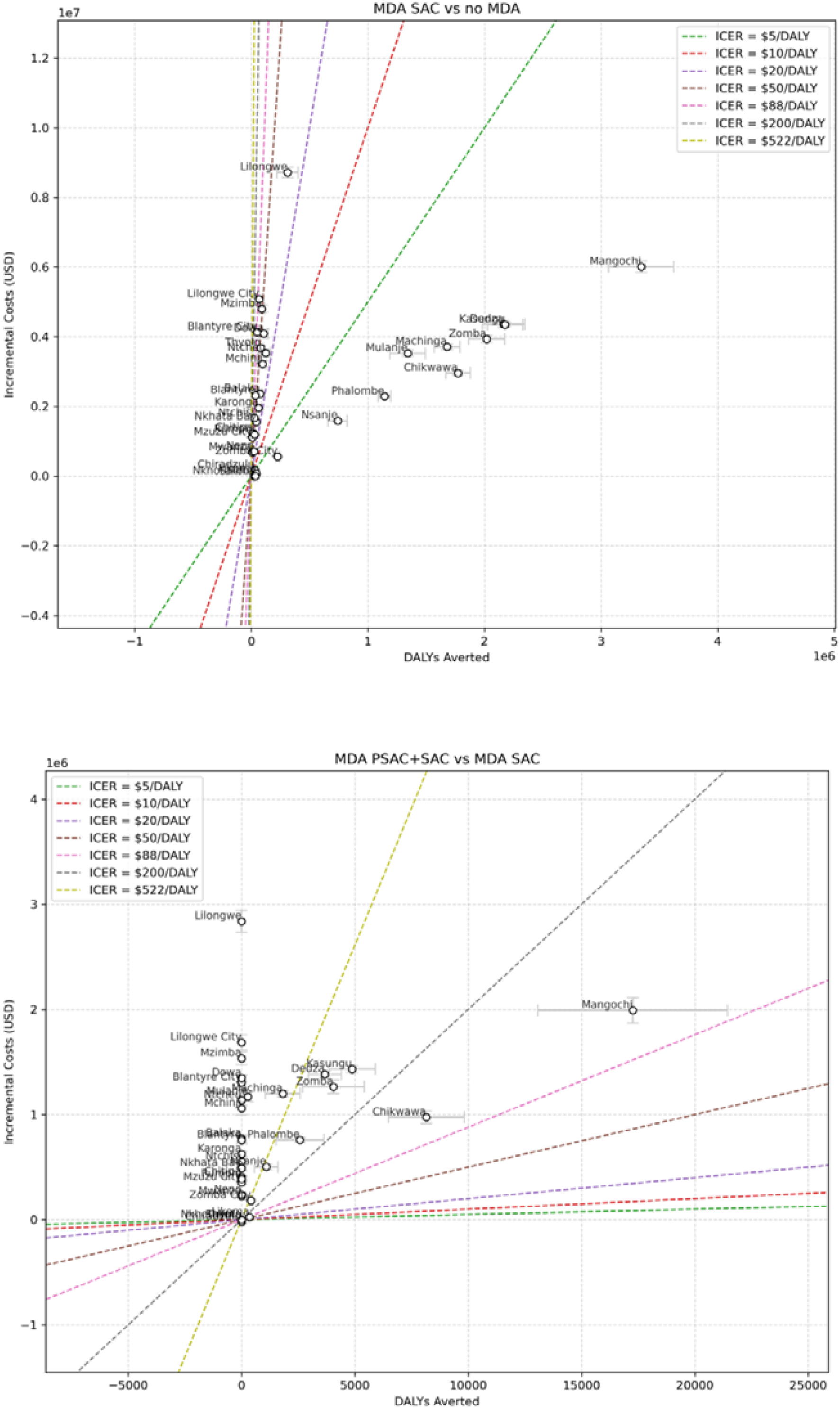
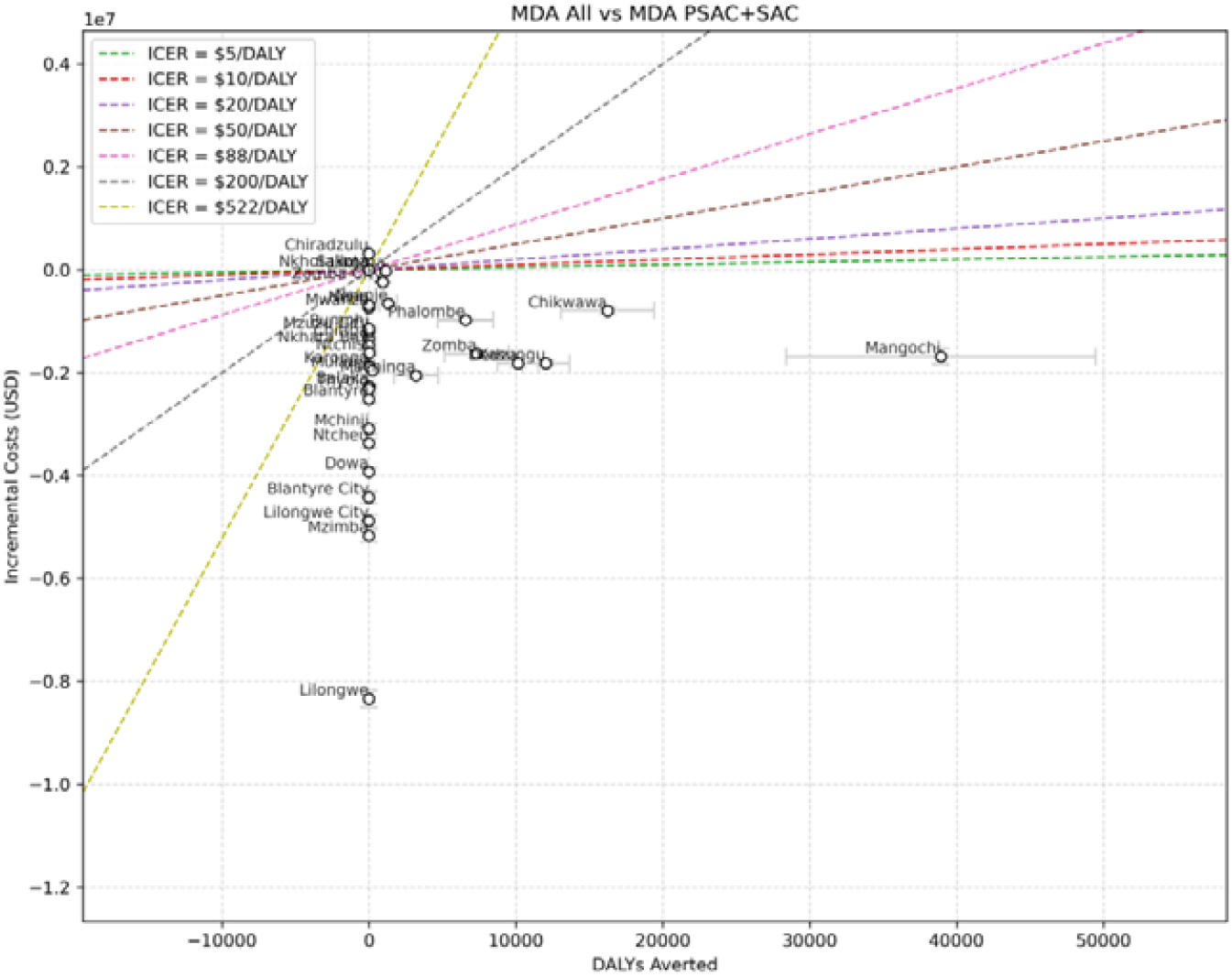
Incremental financial costs versus DALYs averted for MDA SAC, MDA PSAC+SAC, and MDA All for each district. *The panels present the estimated health gains against the financial costs incurred for each delivery strategy. Reference cost-effectiveness thresholds (USD per DALY averted) are included to aid interpretation. Confidence in both costs (vertical bars) and health benefit (horizontal bars) is shown*.

**Figure 4.**
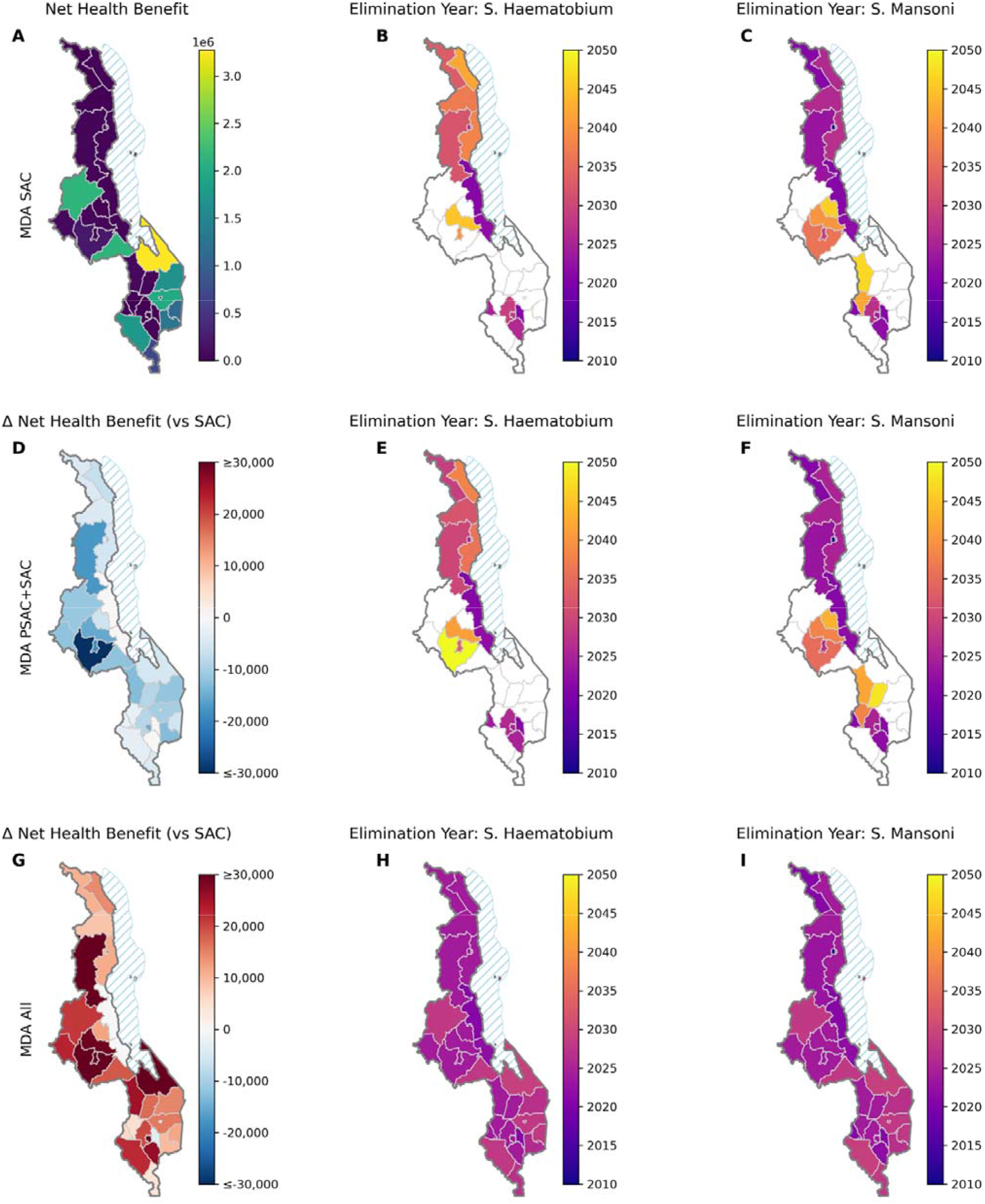
Spatial distribution of net health benefit and elimination timelines under alternative mass drug administration (MDA) strategies in Malawi. Rows show treatment strategies: MDA in school-aged children only (SAC; A–C), preschool- and school-aged children (PSAC+SAC; D–F), and community-wide treatment (All; G–I). Left column shows net health benefit (NHB): absolute NHB for MDA SAC (A), incremental NHB (ΔNHB) for expanded strategies: MDA PSAC+SAC versus SAC (D), and MDA All versus SAC (G). Middle and right columns show predicted elimination year for *S. haematobium* (B, E, H) and *S. mansoni* (C, F, I), respectively. If unshaded, elimination did not occur before 2050.

Expansion from SAC to PSAC+SAC was rarely cost-effective. The national MDA PSAC+SAC ICER was 606 USD/DALY (95% CI 472-695). At district level, PSAC expansion was dominated in 22/32 districts (no incremental health benefit in some runs) and was generally not cost-effective elsewhere. PSAC expansion was only cost-effective in Likoma (ICER 70.8 USD/DALY, 95% CI 51.8-87.3) (Figure 3, Figure 4D).

Community-wide MDA produced a qualitatively different pattern as elimination led to earlier stopping of campaigns. Nationally, MDA All was cost-saving relative to PSAC+SAC but with substantial heterogeneity between districts: MDA All was cost-saving in 11/32 high prevalence districts (2023 prevalence range 13.7 – 41.5%) and dominated (in >80% of model runs) in 16 other districts, indicating limited incremental health gain from adding adults once paediatric strategies had already reduced transmission in these districts. Further, MDA All frequently achieved the highest NHB across a range of opportunity cost thresholds (80-90% of districts, see Appendix), with MDA SAC preferred mainly at lower willingness-to-pay thresholds. When light-intensity morbidity was included, the number of districts in which MDA All was cost-saving compared with PSAC+SAC rose to 29/32 (Appendix Table 10). Discounting costs or health, or applying reduced implementation costs per MDA treatment produced qualitatively similar findings; MDA SAC remained cost-effective, expansion to PSAC+SAC was cost-effective only in 1-3 districts and MDA All became cost-saving in 10-11 districts.

Maximum allowable delivery costs (MADC) per MDA treatment closely tracked baseline epidemiological burden rather than population size or urban-rural status. High-prevalence districts supported substantially larger per-treatment MAC envelopes across all strategies (~ +25-38 USD per treatment for delivery), while low-prevalence districts had limited headroom (generally <1-3 USD per treatment for delivery in MDA SAC) (Table 1). This gradient with respect to burden persisted across strategies; PSAC expansion typically reduced per-treatment MAC relative to MDA SAC, whereas MDA All produced the largest per-treatment MAC values primarily in the highest-burden districts where elimination-driven gains were greatest. Across districts, MAC values were generally higher under Pause WASH than under Scale-up WASH for MDA SAC and MDA All, reflecting larger incremental gains where transmission remained higher (Appendix Table 8).

**Table 1.**
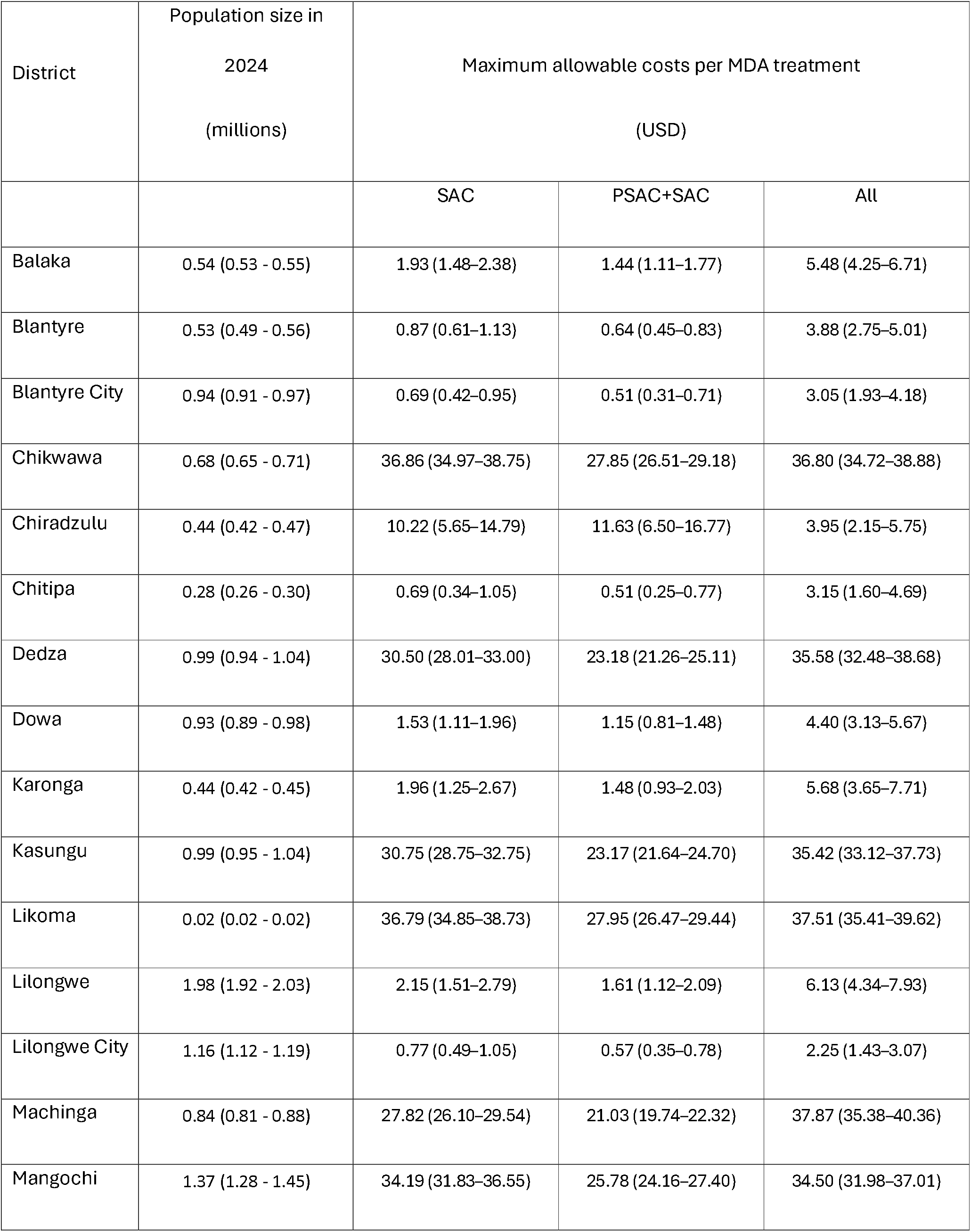
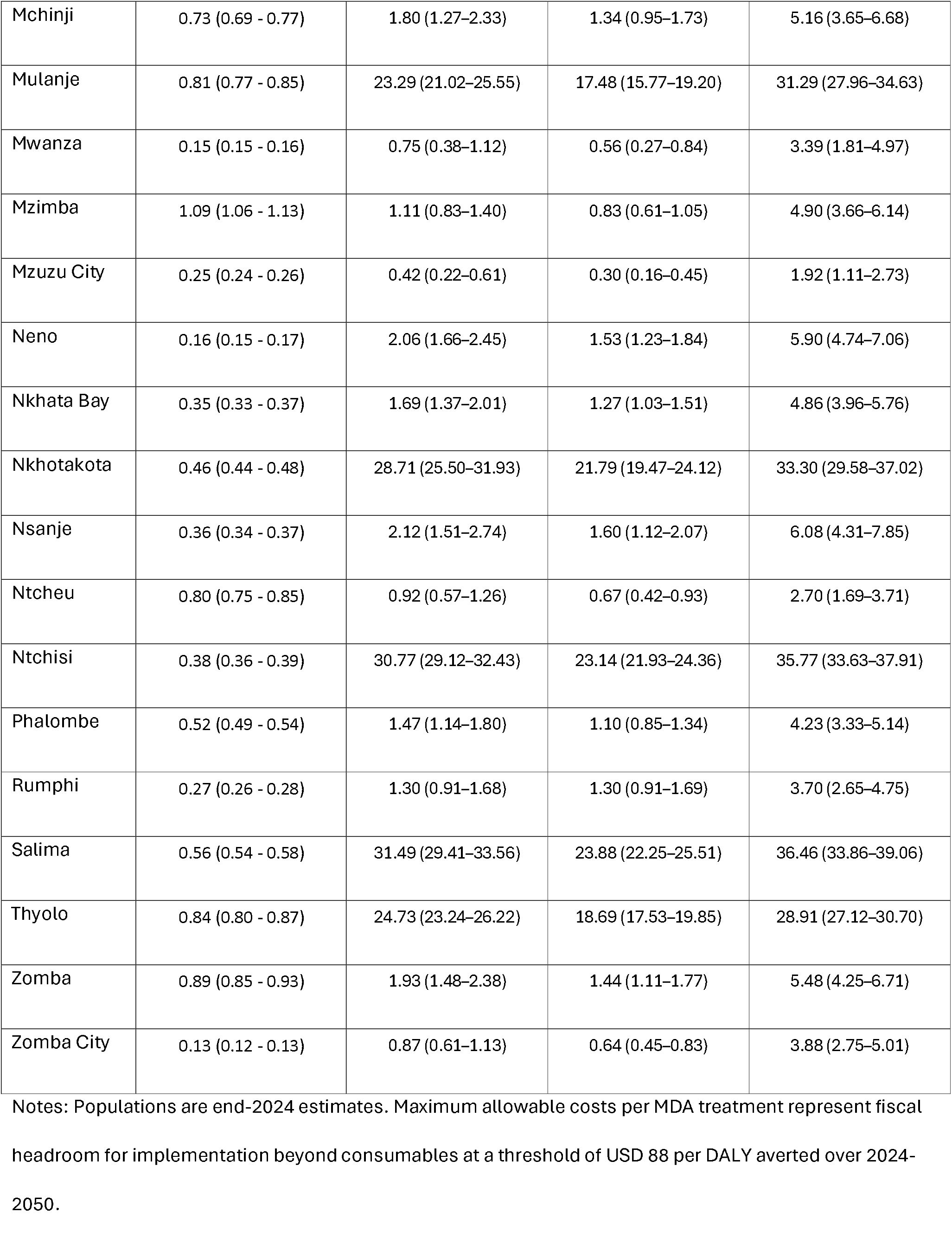
District populations (2024) estimated through modelling and maximum allowable delivery costs (USD) per MDA treatment for MDA strategies.

Including disability for light infections substantially increased MADC. Annualised over 2024-2050, median district MADC increased from 0.25 million USD/year (range 0.03-10.9) to 11.3 million USD/year (range 1.19-347.4) for MDA SAC, with similar increases for MDA PSAC+SAC (median 0.25 to 11.3 million USD/year) and MDA All (median 0.25 to 12.0 million USD/year). The relative distribution across districts was unchanged, with the largest headroom remaining concentrated in high-burden settings.

## Discussion

Community-wide mass drug administration (MDA All) produced the greatest epidemiological impact, consistently driving both *S. haematobium* and *S. mansoni* transmission towards elimination in all districts by 2030. By extending treatment to adults as well as children, this strategy directly targets the adult infection reservoir (~200 million person-years of infection over 2024-2050 compared with ~70 million among school-aged children) which otherwise sustains transmission despite high paediatric coverage.(13) Strategies limited to school-aged children (SAC-only) or SAC plus preschool-aged children (PSAC+SAC), although often cost-effective under constrained thresholds, generally failed to interrupt transmission in high-burden districts. These results are consistent with previous modelling and empirical evidence demonstrating that untreated adult infections maintain substantial reservoirs sustaining transmission, such that paediatric-only strategies reduce morbidity without reliably interrupt transmission.(5, 6, 14, 15)

The greater health impacts of MDA All do not translate into a consistent cost-effectiveness over paediatric strategies. Under a DALY-based evaluation framework, community-wide treatment often appeared either cost-saving or strongly dominated relative to PSAC+SAC. Paediatric treatment captures the majority of measurable morbidity reduction as school-aged children harbour the highest-intensity infections; consequently expansion to adults produces diminishing returns in DALY terms. Where community-wide treatment accelerates elimination sufficiently to shorten programme duration, cumulative costs decline and MDA All becomes economically favourable. Taking into account the long-term economic payoff of achieving elimination (via future avoided treatment and surveillance costs, reduced disability and productivity, avoiding future resurgence risk alongside the health opportunity costs of reallocating resources to other priorities) is crucial – a perspective that can flip the evaluation of community-wide MDA from “too expensive” to cost-saving in the long run. The DALY metric used here, and assigning zero disability weights to light infections, also tends to undervalue the benefit of treating low-intensity infections and averting future infections.(13) Thus, once heavy paediatric infections are managed, further gains from reducing light infections in adults may register as only small DALY improvements. Alternative metrics capturing infection duration or intensity may better reflect the broader impact of transmission reduction. The sensitivity analyses here assigning a small disability weight to light infections shifted community-wide MDA towards cost-saving in more districts, suggesting that the apparent diminishing returns of expansion are partly an artefact of the DALY specification rather than a lack of substantive transmission benefit.

Fiscal constraints, not just epidemiological dynamics, may ultimately dictate strategy choice. Malawi’s national schistosomiasis control budget (approximately ~$100,000 per year) is substantially lower than the projected costs required to cover all at-risk populations.(16) Empirical costing studies indicate higher delivery costs for community-based distribution compared with school-based programmes, reflecting additional workforce and logistical requirements.(17, 18) Even under optimistic assumptions, large-scale expansion to adults would require funding beyond current levels, implying that geographic prioritisation or phased implementation is unavoidable under existing fiscal constraints. Such targeting makes the most of limited resources by prioritizing interventions with the highest immediate health payoff (i.e. treating children in areas of highest risk), but it also means that true elimination, which requires community-wide suppression of transmission, remains out of reach in the short term.

We find here, like others, that community-wide treatment can be cost-effective but economic performance is heterogeneous across the districts of Malawi, where adult infections are frequently light and district-transmission varies widely. (6) We explicitly tie the duration of MDA to achieving local elimination thresholds, allowing us to capture the long-term cost dynamics (e.g. stopping rules once transmission breaks) rather than assuming a fixed-term programme. Our finding that MDA All is “cost-saving” in some districts but dominated in others – was not evident in prior fixed-horizon analyses and reinforces the idea that a short-term increase in spending can pay off by permanently removing the need for ongoing treatment. At the same time, our integrated approach (spanning all districts and both schistosome species) reveals substantial heterogeneity: in some districts with moderate transmission, child-focused MDA nearly achieves local elimination, meaning adult treatment adds little value; in other hotspots, only community-wide coverage achieves control.

WASH improvements reduced exposure gradually but did not alter the relative ranking of MDA strategies.(Grimes, Croll et al. 2014, Grimes, Croll et al. 2015) Recent WHO guidance retains preventive chemotherapy as the core intervention for schistosomiasis, with WASH as a complementary accelerator, however WASH investments deliver important broader health benefits, including reduced risk of diarrhoeal disease, cholera and hepatitis A.(Control of Neglected Tropical Diseases (NTD) 2022)

Several limitations should be considered. Data on baseline prevalence and intensity are sparse in some districts, and these uncertainties propagate through model calibration, potentially affecting local estimates of impact and elimination timeline. Necessary structural simplifications in the transmission model include assumptions around mixing patterns, water contact behaviour, snail ecology and seasonal variation. Assumptions on systematic non-participation are simplified; sustained pockets of untreated individuals can maintain reservoirs of infection, reducing MDA effectiveness and slowing progress towards elimination. Economic conclusions depend on implementation costs, opportunity-cost thresholds and disability weight choices which have been varied and explored. We present maximum allowable costs to show the potential headroom for these costs with each MDA strategy. The health-sector perspective excludes broader societal benefits such as productivity or educational gains, likely rendering programmes conservative in value terms.

Our findings in the Malawian context, characterised by heterogeneous economic value and the need for geographically targeted expansion, align with geostatistical evidence of clustered risk and the sensitivity of programme requirements to spatial scale.(Schur, Vounatsou et al. 2012, Lai, Biedermann et al. 2015) More fundamentally, they highlight a structural tension between strategies that maximise short-term morbidity reduction, as captured by DALY-based evaluation, and those required to achieve transmission interruption over longer time horizons. Because schistosomiasis burden is concentrated in high-intensity infections among children while transmission persistence is sustained by a large reservoir of predominantly low-intensity adult infection, strategies optimised for morbidity control may differ from those required for elimination. Integrating district-level transmission modelling with opportunity-cost–based economic evaluation helps reconcile these perspectives, by explicitly incorporating temporal horizons and the distinct programme objectives when defining efficient pathways for schistosomiasis control and elimination.

## Data Availability

All data produced in the present study are available upon reasonable request to the authors

https://github.com/UCL/TLOmodel.git

## Ethical approval

The Thanzi La Mawa project received ethical approval from the College of Medicine Malawi Research Ethics Committee (COMREC, P.09/23-0297) for the use of publicly accessible and anonymised secondary data. No data were used requiring individual informed consent.

## Data Availability and Code Transparency

All associated code is open-source and available at https://github.com/UCL/TLOmodel/releases. Detailed model documentation, including parameter values, appointment footprints, and calibration methods, is available at Hallett *et al*^25^.

## Acknowledgements

We gratefully acknowledge the guidance, technical input and continued collaboration of the Department of Schistosomiasis and Neglected Tropical Diseases, Ministry of Health, Malawi, whose programme insights and operational expertise were invaluable to this work. We also thank colleagues at the Health Economics and Policy Unit (HEPU) for constructive feedback on the economic evaluation and interpretation of findings.

