## Supplementary material for "Linking transmission dynamics and economic evaluation to assess schistosomiasis programme strategies across districts in Malawi"

### A. Health system model

##### Overview

The schistosomiasis model incorporates a sequence of Health-System Interaction (HSI) processes that determine how individuals with clinically relevant infections engage with diagnosis, treatment, and follow-up care. These HSIs act as the bridge between the natural history of infection and the capacities and constraints of personnel, diagnostics and consumables across each level of Malawi’s health services (community, outpatient and hospitals). The structure ensures that modelled outcomes reflect not only biological processes but also the operational realities of clinical assessment, diagnostic performance, drug availability, and broader health-system bottlenecks.

##### Triggering of Clinical Interactions

Care-seeking behaviour is symptom-driven and probabilistic, reflecting empirical estimates from the 2016 Malawi Integrated Household Survey.(National Statistical Office 2017, Ng'ambi, Mangal et al. 2020, Ng’ambi, Mangal et al. 2020)*.* Individuals enter the HSI pathway when the schistosomiasis module generates a care-seeking event derived from infection severity. Rather than simulating explicit symptoms such as haematuria or abdominal pain, the model uses internal severity indicators (e.g., light, moderate, heavy), which summarise worm burden and associated morbidity. Individuals with moderate or heavy severity are eligible to seek care, with probability of seeking care governed by severity-specific parameters. Those with light-intensity infections are assumed not to seek care specifically for schistosomiasis – although they can seek care for any other conditions.

##### Diagnostic Assessment and Performance of Tests

Once an individual seeks care in the model, a diagnostic HSI is triggered. The symptoms they present with determine which diagnostic test is requested. Symptoms indicative of *S. haematobium* infection (i.e. visible or reported blood in urine) prompt the healthcare worker to request a urine filtration test. In the absence of urinary symptoms, the presentation is treated as potentially due to *S. mansoni* and a Kato-Katz thick smear is requested. This approach mirrors Malawi’s clinical guidelines.(Ministry of Health (MOH) 2023)

Diagnostic test performance is determined by sensitivity and specificity, which depend on the current infection intensity. These values reflect empirical evidence that both urine filtration and Kato–Katz perform well at detecting moderate and heavy worm burdens but have very low sensitivity for low-intensity infections.(Colley, Binder et al. 2013, Bärenbold, Raso et al. 2017, Midzi, Bärenbold et al. 2020) As a result, individuals with light infections frequently test negative, whereas true cases with moderate or heavy infections are more reliably detected. Diagnostic capacity is assumed to be available for individuals eligible for testing, and there is no clinician-level misclassification beyond the probabilistic performance of the tests themselves. A positive diagnostic result leads directly to a treatment referral, while false negatives remain untreated and continue disease progression.

##### Treatment Allocation and Commodity Constraints

Successful delivery of praziquantel depends on two system-level constraints, the availability of sufficient stock at the facility and available patient-facing time of the appropriate cadre of healthcare worker. Individuals who are correctly diagnosed can therefore still fail to receive treatment if either constraint is binding. In this case, they may return to the facility for a repeat appointment or disengage from care.

Routine care requires both laboratory time for diagnosis and clinical time for treatment provision. Praziquantel is administered at age-appropriate doses – 20 mg/kg for children under five years and 40 mg/kg for individuals aged five years and above – using age-specific average weights to determine the required quantity. The efficacy of praziquantel treatment varies between species, with 86.3% adult worm clearance rates for S. mansoni and 94.1% for S. haematobium.(Zwang and Olliaro 2014)

##### Progression to Sequelae and Cross-Module Interactions

A subset of individuals develop chronic complications, most notably bladder cancer (a five-fold increase in bladder cancer risk with S. haematobium infection) and HIV 2.3-fold increase in risk for females with high-intensity S. haematobium infection).(Parkin, Hämmerl et al. 2020, Patel, Rose et al. 2021) When an individual develops another health condition, such as HIV, they would follow the clinical pathway defined in the corresponding disease module according to the National Guidelines. Each pathway includes its own rules for diagnostic assessment and treatment, including the probabilities of referral to higher-level facilities, diagnostic accuracy for chronic pathology, and the availability of specialised services such as surgery, oncology, or palliative care. The probability that an individual receives definitive treatment therefore reflects not only disease severity but also the wider national capacity for managing chronic complications. This ensures that the long-term morbidity and DALY burden in the model correctly incorporate limitations in Malawi’s cancer and surgical care pathways.

##### Mass Drug Administration

MDA operates through a separate delivery mechanism from routine facility-based care but uses the same treatment efficacy parameters as routine case management. Eligibility for each campaign is defined by age-targeting rules: school-aged children (SAC, 5-14 years), pre-school aged children (PSAC, 2-4 years) or adults (15 years and over).Praziquantel dosing is the same as for treatment, using age-group mean body weights to calculate the required number of tablets for each age category.

MDA is assumed to be delivered via a mix of community volunteers and government staff (see Appendix Table 4). No diagnostic testing is undertaken during the MDA campaigns and all eligible individuals receiving treatment through MDA experience the same probabilistic reduction in worm burden as under routine case management.

##### Water, Sanitation, and Hygiene (WASH) Interventions

The TLO model represents three WASH attributes at the individual level: unimproved sanitation, no clean drinking water, and no access to a hand-washing facility. WASH status is initialised at the start of the simulation (1 January 2010) using prevalence distributions derived from the Malawi DHS 2010, stratified by urban–rural residence and, for hand-washing, household wealth quintile.(National Statistical Office (NSO) [Malawi] ICF Macro 2011) Urban–rural status and wealth are assigned first, and WASH properties are then sampled independently using strata-specific probabilities defined in the module resource file:

- urban and rural baseline probabilities for unimproved sanitation;
- urban and rural baseline probabilities for no clean drinking water;
- a baseline probability of no hand-washing access for the lowest wealth quintile, with an odds-ratio applied for each step in wealth to reproduce the observed gradient.

These DHS-based probabilities serve as the model’s initial conditions; DHS 2015 values in the documentation are used purely for external validation and not for parameterisation.(National Statistical Office (NSO) [Malawi] and ICF 2017) After baseline assignment, WASH attributes evolve through fixed transition processes applied every three months. For each attribute, individuals without access face a constant probability of gaining access to improved sanitation, clean drinking water and hand-washing facilities. These transition rates are homogeneous over time and independent across individuals, except for hand-washing access, where transition probabilities incorporate wealth-dependent modifiers to reflect higher uptake in richer households. Transitions operate only in the direction of improvement; once an individual gains access to improved sanitation, clean water, or a hand-washing facility, this status is retained for the remainder of the simulation.

At each simulated year, individual WASH states are aggregated by district to obtain the district-level proportions of unimproved sanitation, lack of clean water, and lack of hand-washing facilities presented in this study (Appendix Figure 1).

We model WASH interventions here as a composite improvement in sanitation and access to clean water. Evidence from Grimes et al. 2014 shows both components reduce exposure to schistosome-containing water, with pooled odds ratios of roughly 0.53 – 0.57 for safe water and 0.59 – 0.69 for adequate sanitation across *S. mansoni* and *S. haematobium*.(Grimes, Croll et al. 2014) To avoid double-counting these overlapping effects, the model applies a single combined reduction in susceptibility of 40% to all individuals, across all age-groups and for both species. In addition to lowering schistosomiasis risk, WASH improvements also reduce the incidence of childhood diarrhoeal disease, reducing the probability of stunting through the relevant TLO modules.

Table 1. Table of parameters

| **Definition** | **Value**  ***S. mansoni*** | **Value**  ***S. haematobium*** | **Notes** |
| --- | --- | --- | --- |
| Reproduction number (R0)  of each species prior  to MDA | 1.66 | 1.6 | (Truscott, Gurarie et al. 2017)  (Turner, Truscott et al. 2017) |
| Harbouring rate (k) | 0.24 | 0.24 | Gamma distribution with shape parameter k.  (Chan, Guyatt et al. 1995) |
| Proportion of the population in each district susceptible to schistosomiasis infection | Calibrated by district | Calibrated by district | See section C on Calibration. |
| Contact rate / exposure rate (β) | PSAC=0.3; SAC=1;  adult=0.05 | PSAC=0.3; SAC=1;  adult=0.05 | (Turner, Truscott et al. 2017) |
| Density-dependent worm fecundity | 0.0006 | 0.0006 | (Anderson, Turner et al. 2016) |
| The time taken for cercariae to develop into mature worms | 25-30 days | 25-30 days | Anderson & May 1992 |
| Adult  Worm lifespan | 5.7 years | 4.0 years | (Turner, Truscott et al. 2017)  (IARC Working Group on the Evaluation of Carcinogenic Risks to Humans 2012)  (Anderson, Turner et al. 2016)  (Fulford, Butterworth et al. 1995) |
| The worm burden threshold denoting a light intensity infection | *<*8 worms | *<*10 worms | Modified from WHO threshold, see Section B – Infection intensity thresholds |
| The worm burden threshold denoting a moderate intensity infection | 8-28 worms | 10-20 worms | Modified from WHO threshold, see Section B – Infection intensity thresholds |
| The worm burden threshold denoting a high intensity infection | *>*28 worms | *>*20 worms | Modified from WHO threshold, see Section B – Infection intensity thresholds |
| The worm burden threshold denoting a high intensity infection in PSAC | 14 worms | 10 worms | Assumed to be half that of older children/adults |
| Efficacy of praziquantel in clearing worm burden | 86.3% | 94.1% | Same efficacy for both MDA and treatment  (Zwang and Olliaro 2014) |
| Proportional reduction in susceptibility to infection when WASH activities are implemented | OR = 0.6 | OR = 0.6 | Composite WASH effect applied as a 40% reduction in susceptibility (OR = 0.6), drawing on Grimes et al. (2014). Evidence indicates similar protective effects across ages: safe water reduces odds of infection (haematobium OR ≈ 0.57; mansoni OR ≈ 0.53) and adequate sanitation provides comparable protection (OR ≈ 0.59–0.69). |

Table 2. ESPEN district-level endemicity and MDA coverage

| District | Endemicity | MDA scheme | School-aged children | | Adults | |
| --- | --- | --- | --- | --- | --- | --- |
|  |  |  | **Reported population requiring treatment** | **coverage** | **Reported population requiring treatment** | **coverage** |
| Dedza | High prevalence (50% and above) | Not delivered | 257879 | 0 | 525392 | 0 |
| Dowa | Moderate prevalence (10%-49%) | PZQ+ALB/MBD | 120644 | 0.190 | 98319 | 0.001 |
| Kasungu | High prevalence (50% and above) | PZQ+ALB/MBD | 262251 | 0.025 | 534301 | 0 |
| Lilongwe | Moderate prevalence (10%-49%) | PZQ+ALB/MBD | 411811 | 0.634 | 335603 | 0.035 |
| Mchinji | Moderate prevalence (10%-49%) | PZQ+ALB/MBD | 93328 | 0.405 | 76057 | 0.001 |
| Nkhotakota | Moderate prevalence (10%-49%) | PZQ+ALB/MBD | 61344 | 0.338 | 49992 | 0.690 |
| Ntcheu | Moderate prevalence (10%-49%) | PZQ+ALB/MBD | 103404 | 0.399 | 84269 | 0.001 |
| Ntchisi | Moderate prevalence (10%-49%) | PZQ+ALB/MBD | 49898 | 1.051 | 40665 | 0 |
| Salima | Moderate prevalence (10%-49%) | PZQ+ALB/MBD | 75134 | 0.735 | 61230 | 1.443 |
| Likoma | High prevalence (50% and above) | PZQ+ALB/MBD | 4528 | 0.794 | 9226 | 0.005 |
| Chitipa | Moderate prevalence (10%-49%) | PZQ+ALB/MBD | 36473 | 1.050 | 29724 | 0 |
| Karonga | Moderate prevalence (10%-49%) | PZQ+ALB/MBD | 57114 | 0.617 | 46544 | 0.017 |
| Nkhata Bay | Moderate prevalence (10%-49%) | PZQ+ALB/MBD | 44370 | 0.711 | 36159 | 0 |
| Rumphi | Moderate prevalence (10%-49%) | PZQ+ALB/MBD | 35786 | 0.279 | 29163 | 0 |
| Balaka | Moderate prevalence (10%-49%) | PZQ+ALB/MBD | 68590 | 0.173 | 55897 | 0 |
| Blantyre | Moderate prevalence (10%-49%) | PZQ+ALB/MBD | 192788 | 1.123 | 157111 | 0 |
| Chikwawa | High prevalence (50% and above) | Not delivered | 174316 | 0 | 355144 | 0 |
| Chiradzulu | Moderate prevalence (10%-49%) | Not delivered | 54654 | 0 | 44540 | 0 |
| Machinga | High prevalence (50% and above) | Not delivered | 233271 | 0 | 475257 | 0 |
| Mangochi | High prevalence (50% and above) | PZQ+ALB/MBD | 362223 | 0.097 | 737979 | 0.068 |
| Mulanje | High prevalence (50% and above) | Not delivered | 211594 | 0 | 431093 | 0 |
| Mwanza | Moderate prevalence (10%-49%) | PZQ+ALB/MBD | 21336 | 1.791 | 17387 | 0 |
| Neno | Moderate prevalence (10%-49%) | Not delivered | 20230 | 0 | 16486 | 0 |
| Nsanje | High prevalence (50% and above) | PZQ+ALB/MBD | 91633 | 0.133 | 186689 | 0.090 |
| Phalombe | High prevalence (50% and above) | PZQ+ALB/MBD | 133866 | 0.514 | 272734 | 0 |
| Thyolo | Moderate prevalence (10%-49%) | Not delivered | 110272 | 0 | 89865 | 0 |
| Zomba | High prevalence (50% and above) | Not delivered | 262928 | 0 | 535679 | 0 |
| Mzimba | Moderate prevalence (10%-49%) | PZQ+ALB/MBD | 180903 | 1.389 | 147426 | 0.001 |
| Lilongwe City | Moderate prevalence (10%-49%) | PZQ+ALB/MBD | 411811 | 0.634 | 335603 | 0.035 |
| Blantyre City | Moderate prevalence (10%-49%) | PZQ+ALB/MBD | 192788 | 1.123 | 157111 | 0 |
| Mzuzu City | Moderate prevalence (10%-49%) | PZQ+ALB/MBD | 180903 | 1.389 | 147426 | 0.001 |
| Zomba City | High prevalence (50% and above) | Not delivered | 262928 | 0 | 535679 | 0 |

Reported schistosomiasis endemicity classifications and MDA coverage by district for school-aged children and adults, based on 2022 ESPEN data.(World Health Organization Regional Office for Africa 2024) “Population requiring treatment” reflects ESPEN estimates of the target population for each age group under Malawi’s MDA scheme. Coverage denotes the proportion of the eligible population reported to have received praziquantel during the 2022 campaign. Abbreviations: PZQ praziquantel; ALB albendazole; MBD mebendazole.

### B. Schistosomiasis transmission model

The schistosomiasis model explicitly tracks individual worm burden for two species of schistosomes, *S. mansoni* and *S. haematobium*. Transmission dynamics are modelled at the district level, with no assumed population movement between districts. The basic reproduction number (*R0*) is species-specific and fixed, this is used as a transmission coefficient encapsulating ecological and behavioural determinants of transmission (snail population, environmental survival of free-living stages, and human–water contact). The proportion of the population susceptible to infection is calibrated to existing epidemiological data.

Age-specific exposure rates (*β_j_*) drive both infection and onwards transmission, with individual harbouring rates drawn from a Gamma distribution reflecting the clustering of worms in high-risk individuals. The probability of infection for an individual is determined by their susceptibility, adjusted by the harbouring and exposure rates, and by the aggregate worm burden of that species in the district.

For each species of *Schistosoma* (*d*), the following infection process occurs each month.

The district-level reservoir of infection is defined as the total worm burden harboured by individuals in the district adjusted by age-dependent exposure. Upon infection, newly acquired worm burdens are sampled from a Poisson distribution, where the rate parameter depends on the district-level reservoir, individual harbouring rates, and contact rates.

The establishment of new worms is density-dependent, limited by the existing worm burden within the host and species-specific worm fecundity. Once established, worms mature within 30–55 days. Maturation renders the host infectious, as worms pair and begin producing eggs.

##### Definitions

Let *i* index districts, *j* age groups, and *k* individuals.

*N_i,j_*: Number of individuals in district *i* and age group *j*

*B_i,j,k_*: Worm burden for individual *k* in district *i* and age group *j*

*β_j_*: Age-specific exposure rate for age group *j*

*N_i_=∑_j_ N_i,j_*: Total population in district *i*

The mean worm burden in district *i*, age group *j* is:

$$\bar{B}_{ij}=\left( 1/N_{ij} \right)\cdot\sum_{k\in(individuals in i,j)} B_{ijk}$$

where *k* ∈ *(i, j)* indicates individuals in district *i* and age group *j*.

The district-level reservoir (summing contributions across all age groups) is calculated using the contribution of age group *j* to the reservoir in district *i*, weighted by the population size and exposure rate. Summing across all age-groups gives the district reservoir:

$$R_{i}=\sum_{j} \beta_{j}\cdot\bar{B}_{ij}\cdot\frac{N_{ij}}{N_{i}}$$

##### Exogenous reservoir of infection

In addition to the district-level reservoir above, we applied an exogenous prevalence-based adjustment to prevent spurious “cliff-edge” declines in prevalence after intensive intervention rounds, whilst preserving the potential for elimination at genuinely low levels of transmission.

At each time step *t*, the national prevalence of infection, *p_t_*, was computed as the proportion of living individuals harbouring a non-zero worm burden, constrained to the interval [0,1]. A reference prevalence, *p_ref_*, was fixed from the baseline national prevalence.

A unitless taper function was then defined as

$$s_{t}=min\left\{ 1, \left( \frac{p_{t}}{p_{ref}} \right)^{\gamma} \right\}$$

with γ determining the rate at which the adjustment vanishes as national prevalence declines. The taper was applied multiplicatively to the reservoir in each district:

Let *ER_i,t_* denote the human reservoir (worm-equivalents) in district *i* at time *t*. We define an exogenous reservoir:

$$R_{i,t}^{adj}=R_{i,t}\left( 1+b_{rel}s_{t} \right)$$

Where *R_i,t_* is the summed, age-weighted worm burden in district *i* at time *t.* For the analyses presented here, we set *b_rel_* =0.02 and γ=1.3. With these values, the adjustment increases the reservoir by approximately 2% at baseline, ~1.9% when national prevalence halves, and falls to ~0.2% when national prevalence is ~1% of baseline. The correction thus fades naturally as prevalence approaches zero, ensuring that elimination remains possible without imposing an artificial floor.

The adjustment was applied after aggregation of individual burdens to district reservoirs and before mapping to the force of infection (FOI). This preserves the biological interpretation of the reservoir: *R_adj_* remains in worm-equivalents and reflects a residual, nationally scaled contamination pressure.

##### Acquisition of new worms

At baseline, the proportion of individuals susceptible to infection in each district was calibrated to reproduce observed prevalence among school-aged children. For each district, the required number of susceptible individuals was first allocated to those without improved sanitation, the remaining susceptible quota was randomly assigned among those with sanitation. This effectively links individual access to safe water and sanitation to infection risk, ensuring district-level prevalence levels are aligned with data.

Each individual’s susceptibility status (*s_k_*) was thereafter treated as fixed, except in cases where WASH improvements occurred during the simulation. In such cases, individuals newly gaining improved sanitation experienced a reduced probability of remaining susceptible, with susceptibility removed in a proportion of cases determined by the WASH intervention effect.

Each individual is randomly assigned a rate of harbouring worms (*h_k_*) drawn from a gamma distribution. This reflects clustering of worms in high-risk individuals and is not age-specific. Individual harbouring rates are multiplied by age-specific exposure and susceptibility (calibrated, see details below) to produce effective rates of worm harbouring (ϕ):

$$\varphi_{k}=h_{k} \cdot\beta_{j}\cdot s_{k}$$

The district-level reservoir *R_adj_* calculated above is a proxy for egg output and thus for contamination pressure. Eggs themselves do not establish new infections directly: they must pass through several stochastic processes - miracidia infecting snails, development into cercariae, and subsequent human exposure - each with low and context-dependent probability of success. To map the contamination proxy to an effective force of infection (FOI), we applied a fitted species-specific transmission coefficient, denoted *κ_d_*:

$$\lambda_{i,t}=\kappa_{d}\cdot R_{i,t}^{adj}$$

Here *κ_d_* is represented by an “*R0*” parameter for each district. This should not be interpreted as the epidemiological basic reproduction number. Instead, it is a calibrated linear multiplier that encapsulates ecological and behavioural determinants of transmission (snail population competence, environmental survival of free-living stages, and human–water contact). This formulation ensures that *λ_i,t_* represents the expected rate of new worm establishment events arising from the prevailing contamination pressure in district *i*.

The number of newly infecting worms in the individual is sampled from a Poisson distribution:

$$Newly acquired worms \sim Poisson\left( \lambda=W_{i} \cdot\varphi_{k} \right)$$

The probability of establishment of newly infecting worms decreases with the individual’s existing worm burden (*B_k_*) and depends on species-specific worm fecundity (*f_s_*):

$${probability of infection}_{k}=\left\{ \begin{matrix} 1, if U_{k}<\exp\left( -B_{k} \cdot f_{s} \right) \\ 0, otherwise \end{matrix} \right.$$

where *U_i_* ∼ Uniform(0*,*1) is a random sample.

Newly established worms mature after a random delay between 30 and 55 days. This stochastic process ensures that each host’s ability to harbour new worms is probabilistically determined based on current infection status and the biological constraints of worm reproduction.

##### Initialising the simulation

At the start of the simulation (year 2010), we initialise the infection states for individuals within each district and species by distributing worms across individuals according to their calibrated harbouring rates and age-specific contact patterns.

For district *i*, the initial reservoir of worms is defined as:

$$R_{i}=N_{i}\cdot{MWB}_{2010,i}$$

where *N_i_* is the population size and *MWB_2010,i_* is the mean worm burden assigned for initialisation. Worms are allocated randomly to individuals within each district. First, each person’s contact and susceptibility factor is determined by multiplying their calibrated susceptibility [0,1] with the age-specific exposure weight *β _j_*, where $j (PSAC, SAC, Adults)$. These factors are then combined with each individual’s gamma-distributed harbouring rate, producing an overall acquisition rate. Worms are assigned proportionally to these rates by random sampling without replacement, such that individuals with higher harbouring and exposure and more likely to be allocated greater worm burdens.

This procedure is repeated independently for *S. mansoni* and *S. haematobium* in each district. All natural history parameters, including species-specific transmission coefficients and worm lifespan etc, are held constant. Only the initial state (district and age-specific mean worm burdens) is set from the 2022 distribution.

##### Infection intensity thresholds

Clinical classification of schistosomiasis is commonly based on infection intensity, defined by egg counts in urine or stool. According to World Health Organization (WHO) guidelines, a heavy-intensity infection is reported when egg excretion exceeds 50 eggs per 10 ml of urine for *Schistosoma haematobium* or 400 eggs per gram (EPG) of stool for *Schistosoma mansoni*. Moderate-intensity infections are defined as 25–49 eggs per 10 ml for *S. haematobium* and 100–399 EPG for *S. mansoni*, with lower values classified as light-intensity infections.

As our model tracks the worm burden of each individual, we converted these WHO thresholds into equivalent values of WB, under the assumption of a linear relationship between egg output and worm burden. For a given infection, worm burden can be expressed as:

$$WB=\frac{Excreted eggs}{Eggs per worm pair} \times2$$

where the factor of 2 accounts for the requirement of a mated worm pair for egg production.

##### S. haematobium

For *S. haematobium*, each worm pair produces on average 52 eggs per 10 ml urine (epml) (Truscott et al. 2017).Translating WHO thresholds into worm burden gives:

- Heavy-intensity infection: *WB* ≥ 20 worms
- Moderate-intensity infection: 10 ≤ *WB* ≤ 19 worms

• Light-intensity infection: *WB <* 10 worms

##### S. mansoni

For *S. mansoni*, the expected average daily egg output per worm pair is approximately 100 eggs. To relate this to diagnostic egg counts, we apply the ratio of the Kato–Katz thick smear weight to average daily faecal production, corresponding to ~0.05 eggs per slide or 1.2 eggs per gram (EPG) (Barenbold 2021). This yields the following worm burden thresholds:

- Heavy-intensity infection: *WB* ≥ 28 worms
- Moderate-intensity infection: 7 ≤ *WB* ≤ 27 worms
- Light-intensity infection: *WB <* 7 worms

##### Disability weights by infection intensity

To translate infection states into health loss, we assigned disability weights to each state. We used the maximum disability weight corresponding to the expected spectrum of symptoms present at a given infection intensity. The weights applied were as follows:

Table 3. Disability weights assigned to each infection state.

| Infection state | Disability weight (DW) |
| --- | --- |
| Mild infection | 0.000 |
| Moderate *Schistosoma mansoni* | 0.011 |
| Heavy *S. mansoni* | 0.325 |
| Moderate *Schistosoma haematobium* | 0.011 |
| Heavy *S. haematobium* | 0.325 |

These values reflect the assumption that moderate intensity infections may lead to mild symptomatic burden, whereas heavy infections are associated with substantially greater morbidity.

In our modelling framework, for an individual in a given infection intensity category, the disease‐adjusted life years lost (i.e. years lived with disability, YLD) were computed via:

$${YLD}_{i}=I_{i}\times DW_{i}\times L$$

where $I_{i}$ is the incidence (or prevalence) of infection state $i$, $DW_{i}$ is the corresponding disability weight as above, and $L$is the duration (in years) of that state. In cases where infections progress or regress over time, we apportioned $L$accordingly to the sub‐intervals spent in each intensity category.

We treated the “mild infection” category as having zero disability weight (i.e. no symptomatic burden) (DW = 0.000), based on prior conventions in parasitic disease burden estimation. The choice of the maximum disability weight per class ensures a conservative (upper‐bound) estimate of morbidity burden under the assumption that all expected symptoms for that intensity are represented.

Figure 1. District-level estimates of WASH-related conditions in 2023, before any interventions are enacted.

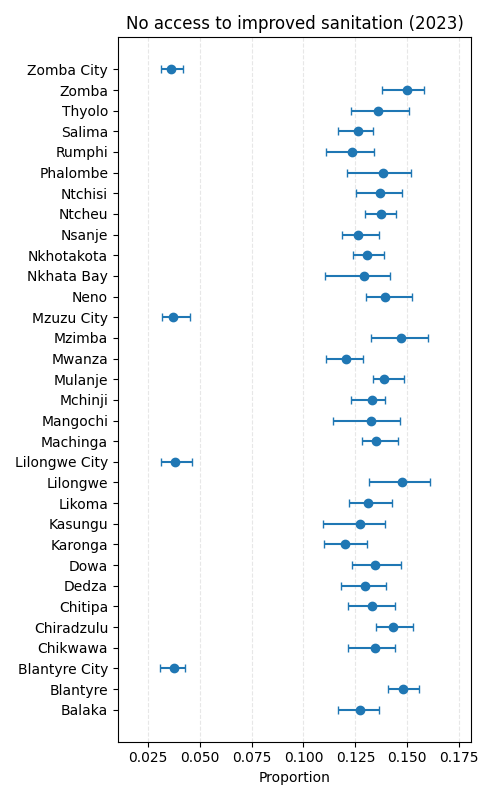

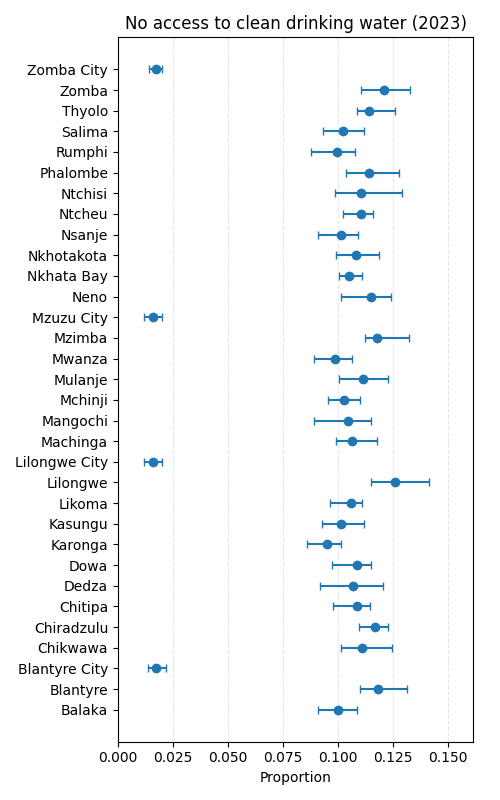

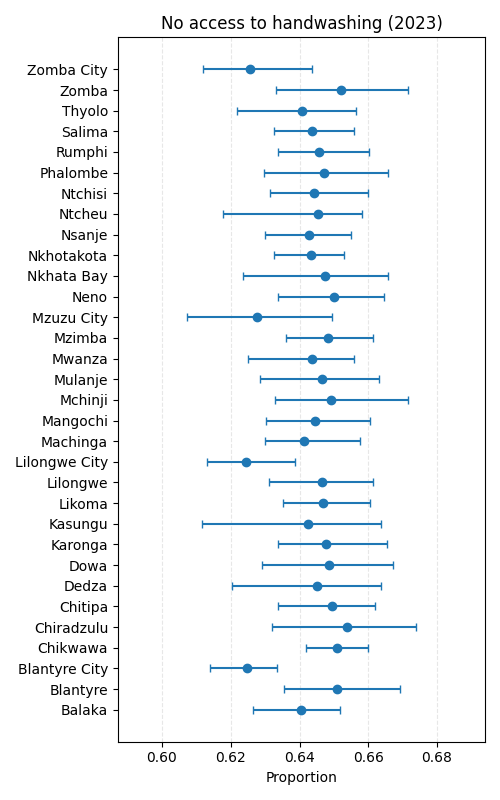

*Each panel presents one WASH property, with points representing the mean estimate across model runs, and horizontal bars indicate the associated 95% uncertainty interval.*

### C. Calibration

The calibration step had two goals: (i) to characterise the relationship between susceptibility and prevalence in a fully susceptible population and (ii) to use this relationship, together with prevalence data, to derive district-level initial conditions (proportion susceptible and mean worm burden) for the simulation.

##### Endemic equilibrium and the susceptibility–prevalence relationship

We first used the TLO framework to determine the endemic equilibrium for *S. mansoni* and *S. haematobium* in a fully susceptible, homogeneously mixing population. A representative Malawi population of 5,000 individuals was simulated, with birth and death rates set equal to maintain a stable population size and continuous replenishment of susceptible individuals. For each species, we ran eight simulations with randomly drawn starting values for mean worm burden and prevalence, and simulated for 100 years with a fixed, literature-based $R_{0}$. No treatment or MDA programmes were enabled.

Endemic equilibrium was defined as the state in which prevalence and mean worm burden stabilised over time. Convergence in school-aged children (SAC) and adults during the final 10 years of the simulation was confirmed by graphical inspection of prevalence and mean worm burden trajectories. For each species, the endemic equilibrium prevalence in a fully susceptible population, $P_{fully susceptible}$, was then calculated as the average monthly prevalence across all runs during 2090–2099. This yielded equilibrium prevalences of 75.9% for *S. mansoni* and 61.7% for *S. haematobium*.

These equilibrium values provide a mapping between the proportion of the population that is susceptible and the resulting endemic prevalence. In particular, for a district in which the entire population is susceptible, prevalence will tend towards $P_{fully susceptible}$; for districts with lower observed prevalence, the implied proportion susceptible is lower.

##### Using 2022 prevalence to determine the proportion susceptible by district

To set realistic starting conditions for the transmission model, we require district-specific estimates of the proportion of the population susceptible to each species. Although historical prevalence data from Malawi (1982–2015, pre-MDA) exist, they are sparse, geographically patchy and generally biased downwards, particularly around 2010. In contrast, 2022 surveillance and ESPEN prevalence bands provide a more complete and diagnostically robust picture of the contemporary infection landscape.

Because the biological and transmission parameters (including $R_{0}$) are fixed from the literature, medium-term behaviour is driven primarily by the initial infection state. Initialising from low, noisy 2010 prevalence values leads to “cold starts” in which infection collapses under observed SAC-only MDA, inconsistent with empirical persistence. We therefore treat the 2010 infection state as an unobserved latent quantity and use the better-resolved 2022 district-level prevalence to infer it. This is analogous to state initialisation or back-casting in state–space models: the latent state is inferred from a more informative, later snapshot, while the transmission parameters remain fixed.

Let $P_{i}^{(2022)}$ denote the 2022 prevalence for species $s$ in district $i$, derived either from point estimates or from ESPEN prevalence bands. The implied proportion of the population in district $i$ that is susceptible to that species, $S_{i}$, is then:

$$S_{i}=\frac{P_{i}^{(2022)}}{P_{fully susceptible}}$$

Thus, districts with 2022 prevalence close to the endemic equilibrium are inferred to be largely susceptible, whereas districts with lower prevalence are inferred to have a smaller susceptible fraction. These $S_{i}$ values are used to scale susceptibility at the start of the simulation.

##### Deriving district mean worm burden from 2022 prevalence

Given a district-specific prevalence $P_{i}^{(2022)}$, we then derive the corresponding mean worm burden $M_{i}$ using the standard negative-binomial relationship between mean worm burden, aggregation and prevalence. If worms per host follow a negative-binomial distribution with mean $M$ and clumping (aggregation) parameter $k$ (the harbouring rate), the prevalence $P$ is given by:

$$P=1-{(1+\frac{M}{k})}^{-k}.$$

For each district $i$ and species $s$, we invert this expression to obtain the mean worm burden $M_{i}$ compatible with the chosen $k$ and the 2022 prevalence $P_{i}^{(2022)}$. As 2022 prevalence was provided as a band (e.g. 10–49% or ≥50%), we map bands to the mid-range point values and apply the same relationship to obtain $M_{i}$.

Together, the district-specific susceptible fraction $S_{i}$ and mean worm burden $M_{i}$ define the calibrated infection intensity and susceptibility landscape used to initialise the model. These quantities are held fixed when the simulation is rewound to 2010, and the observed 2010–2024 MDA history is then replayed to generate a realistic 2024 baseline. As transmission coefficients remain fixed, this warm-start approach avoids initial-condition artefacts (premature fade-out) inconsistent with empirical studies of persistence that arise if low 2010 values are used directly, and ensures that the model enters the projection window with infection levels consistent with contemporary epidemiology.

#### E. Validation

Model calibration was performed using district-level schistosomiasis prevalence data from 2010, with model parameters adjusted to reproduce the spatial distribution of infection intensity at baseline. To validate model performance independently of the calibration data, we compared modelled prevalence in 2019 with empirical observations from district-level surveys conducted in 2019.

For each district, empirical prevalence was summarised as the observed minimum and maximum values across all survey sites within a district, representing the uncertainty range within which the true district-level prevalence lies. Districts reporting zero infections were represented by a lower and upper bound or zero. In districts where no numerical prevalence was available but a non-zero endemicity classification was provided by ESPEN, these observations were treated as missing data.

Model outputs for 2019 were expressed as point prevalence estimates (mean proportion of school-aged children infected), converted to percentages. The district-level validation plots in which empirical uncertainty ranges were shown against the modelled estimates, demonstrate that simulated 2019 prevalence closely matched empirical data. In the majority of districts with available survey information, model estimates lay within the observed uncertainty ranges. Where model outputs fell marginally outside these intervals, deviations were small in absolute magnitude: the 90^th^ percentile distance between the model estimate and the nearest empirical boundary was 6.0 percentage points for *S. haematobium* and 25.7 percentage points for *S. mansoni*. These differences are consistent with sampling variability in school-level surveys and the expected divergence when comparing district-level modelled estimates against single site surveys.

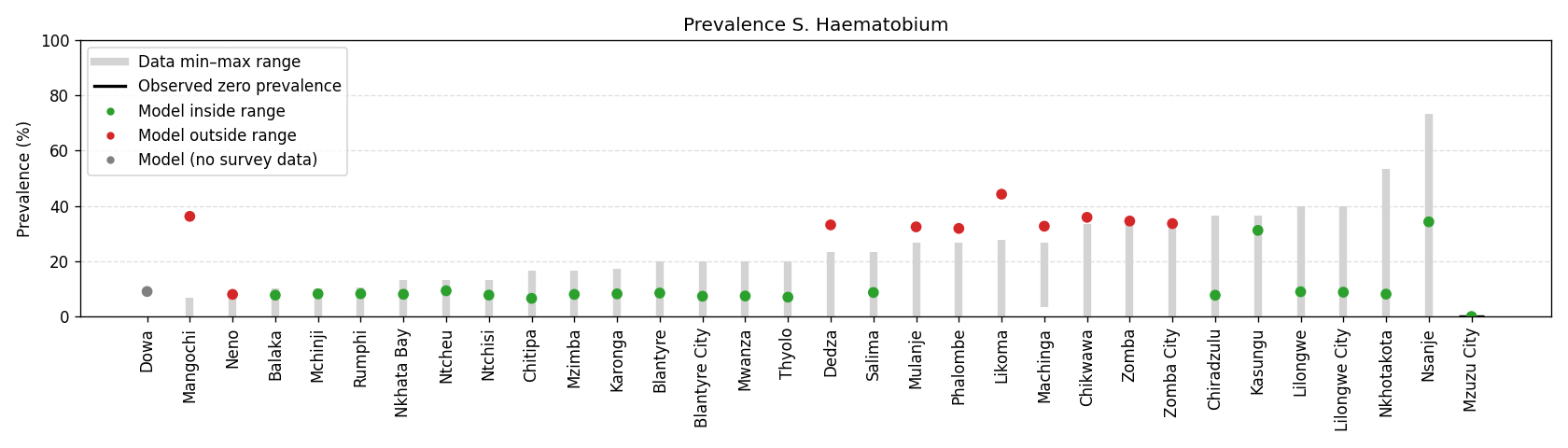

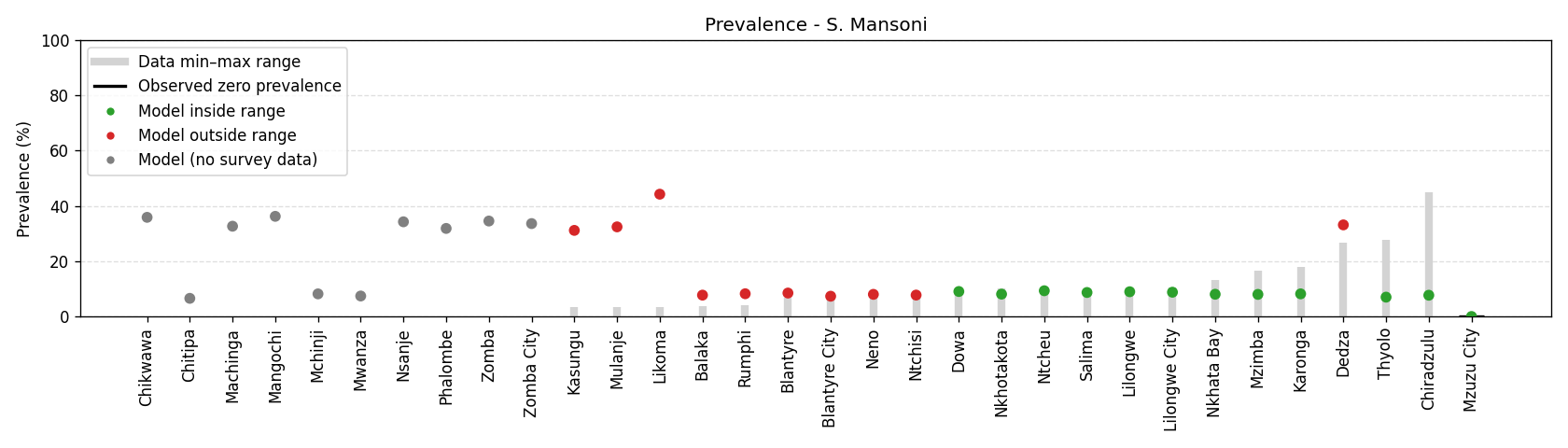

Figure 2. District-level prevalence of S. haematobium and S. mansoni against empirical survey data collected by ESPEN for 2019 – the most recent year in which survey data are available.

*Vertical bars represent the observed uncertainty intervals (minimum – maximum prevalence) in districts where survey data were available. Black horizontal ticks denote districts where surveys reported zero prevalence. Model estimates are shown as: grey in districts without numerical survey data, green when the modelled value falls within the empirical range, red when it lies outside.*

#### F. Costs

##### Updating the Malawi cost-effectiveness threshold to 2025 US$

The cost-effectiveness threshold (CET) was updated from a previously estimated value of US$61 per disability-adjusted life year (DALY) averted expressed in 2018 US dollars to 2025 price levels using a market-consistent approach that preserves the real value of the threshold in local currency before re-expressing it in US dollars.(Ochalek, Lomas et al. 2018) Macroeconomic inputs were taken from World Bank country indicators for Malawi (period averages): (i) the official exchange rate (MWK per US$), and (ii) annual consumer price inflation (CPI, %). Let CET_t,USD_ and CET_t,MWK_ denote the threshold in US dollars and Malawian kwacha (MWK), respectively, in year $t$, and let $XR_{t}$ denote the annual average exchange rate (MWK per US$). The base-year threshold was first converted into local currency according to:

$${CET}_{2018,MWK}={CET}_{2018,USD}\times{XR}_{t}$$

where CET_2018_=61 and XR_2018_=732.3 MWK per US$, giving CET_2018,MWK_=44,670.3 MWK per DALY averted.

To maintain constant real value in local purchasing power, the MWK-denominated threshold was inflated using annual Malawi consumer price index (CPI) inflation rates. Denoting annual inflation by $\pi_{y}$, the updated threshold in 2025 MWK is given by:

$${CET}_{2025,MWK}={CET}_{2018,MWK}\times\prod_{y=2019}^{2025} (1+\pi_{y})$$

Applying the CPI multipliers below gives a CET_2025,MWK_~154,498 MWK per DALY averted. The updated threshold was then re-expressed in USD using the 2025 exchange rate (XR_2025_=1,750.8 MWK per US$.

Inflation, consumer prices

(annual %, period average):

2019: 9.4%

2020: 8.6%

2021: 9.2%

2022: 20.9%

2023: 28.7%

2024: 32.3%

2025: 29.5%

CET_2025,USD_=CET_2025,MWK_ / XR_2025_ ~ 88.24 US$ per DALY averted.

###### Additional threshold (upper bound)

For sensitivity analysis, we also consider an upper-bound CET of US$116 per DALY averted (2016 US$). Applying the same market-consistent conversion and inflation approach yields an equivalent value of approximately US$200 per DALY averted in 2025 US$.

##### MDA programme costs

Cost estimates for MDA were derived using an economic-costing framework that combines programme delivery costs with age-specific praziquantel dosing requirements. All costs are reported in 2025 US dollars.

###### Programme delivery costs

Direct programme costs were taken from published estimates for Malawi, based on four implementation rounds covering approximately 50,000 individuals per round.(Morozoff, Avokpaho et al. 2022) These estimates include routine financial expenditure, routine opportunity costs (time costs for government staff and volunteers), and supportive financial inputs (e.g. supervision, electronic data capture). Appendix Table 4 summarises the mean financial costs per person treated across core programme components, adapted from the primary costing study.

Using the same method as for the CET, and treating costs as 2019 US$, the 2025 US$ inflated equivalents for the implementation costs are:

0.48 (2019 US$) → 0.65 (2025 US$)

These programme-level costs represent the economic cost of delivering an MDA round, encompassing programme management, community sensitisation, training and drug delivery. They were originally estimated for moderately sized campaigns and are not assumed to scale linearly with population size. In the main analysis of the manuscript, these financial costs were incorporated; in the supplementary sensitivity analyses we review the costs for consumables only and consumables plus 0.5 x financial implementation costs.

###### Praziquantel dose and consumables cost

Age-specific dosing requirements were based on standard weight-for-age assumptions for Malawi:

- Preschool-aged children (PSAC; 0–4 years): 17.5 kg, 20 mg/kg dose
- School-aged children (SAC; 5–14 years): 30 kg, 40 mg/kg dose
- Adults (≥15 years): 62 kg, 40 mg/kg dose

The unit cost of praziquantel was taken as USD 0.05 per tablet-equivalent dose for children and USD 0.10 for adults, consistent with procurement prices reported in the literature and aligned with previous cost-effectiveness analyses.

###### Population weighting and mean consumables cost per treatment

To calculate the mean consumables cost for mixed-age strategies, Malawi’s 2024 demographic structure was used:
PSAC = 15.0%; SAC = 22.7%; adults = 62.3%.

A weighted mean cost per treatment in MDA campaigns was therefore:

$$(0.15+0.227)\times0.05 + 0.623\times0.10 = 0.081 USD$$

This average was applied in all scenarios treating the full population aged ≥2 years. Age-restricted strategies used the relevant age-specific unit costs.

###### Application in the model

For each district, the total MDA cost was calculated as:

$$C_{d,s}=N_{d,s}\times c_{a}$$

where $N_{d,s}$ is the number treated in district $d$ under strategy $s$, and $c_{a}$ is the age-appropriate consumables unit cost (USD 0.05, or USD 0.081 for mixed-age scenarios).

These costs were combined with model-estimated DALYs to derive incremental cost-effectiveness ratios and district-level net health benefit. All analyses were conducted over the period 2024–2050 without discounting in the base case.

##### Maximum implementation costs

For each district and MDA strategy, we estimated the maximum implementation costs that could be incurred beyond consumables while remaining cost-effective at a health opportunity cost threshold (*λ*) of USD 88 per DALY averted over 2024-2050. For district *i*, we defined the maximum allowable cost (MAC) as:

$${MAC}_{(i)}=\Delta{DALY}_{i}\cdot\lambda-\Delta C_{i}$$

where *ΔDALY* is the DALYs averted versus the comparator (no MDA) and *ΔC* is the incremental consumables costs for the strategy. By construction, a positive MAC indicates fiscal space for delivery functions such as logistics, staff time, training, social mobilisation, supervision, monitoring and evaluation, while meeting the cost-effectiveness threshold. Non-positive values indicate that, given consumables costs alone, the strategy would not fall below the threshold.

##### Economic evaluation

Cost-effectiveness was assessed using incremental cost-effectiveness ratios (ICERs) and net health benefit (NHB) for every district. ICERs were estimated sequentially: (i) MDA SAC vs no MDA, (ii) PSAC+SAC vs SAC, and (iii) all ages vs PSAC+SAC. NHB was defined as the difference between the health gains of each strategy measured in DALYs averted and the health opportunity cost of its implementation, with programme costs converted to health units using the willingness-to-pay threshold. NHB was calculated relative to no MDA.

$${ICER}_{(A\to B)}=\frac{C_{B}-C_{A}}{E_{B}-E_{A}}$$

where *C* denotes total cost and *E* the total DALYs incurred for strategies A and B.

$${NHB}_{(i)}=\Delta{DALY}_{i}\frac{\Delta C_{i}}{\lambda}$$

All costs and health outcomes were calculated over the period 2024–2050 and initially undiscounted.

Table 4. Unit costs for programme components of schistosomiasis control in Malawi

| **Programme component** | **Cost** |
| --- | --- |
| **Programme management** | $ 0.15 |
| **Community sensitisation** | $ 0.06 |
| **Training** | $ 0.07 |
| **Drug delivery** | $ 0.20 |
| ***Average unit costs‡*** | *$ 0.48* |

*Itemised financial costs for key programme components, expressed in 2022 USD. Routine costs reflect expenditures associated with standard health system delivery, while supportive costs capture additional resources required for programme implementation, including NGO supervision and enhanced community sensitisation activities. Opportunity costs represent the value of staff and volunteer time required for programme delivery. Adapted from Morozoff et al 2022.*(Morozoff, Avokpaho et al. 2022)

#### G. Additional results

Table 5. Person-years infected with any Schistosoma species during 2024-2050 by district and age group (millions).

| District | Age | MDA All | MDA PSAC | MDA SAC | no MDA |
| --- | --- | --- | --- | --- | --- |
| Balaka | **Adults** | 0.03 (0.02–0.03) | 1.02 (0.77–1.28) | 1.07 (0.81–1.32) | 3.65 (3.22–4.18) |
|  | **Infant** | 0.00 (0.00–0.00) | 0.00 (0.00–0.01) | 0.00 (0.00–0.01) | 0.09 (0.08–0.12) |
|  | **PSAC** | 0.00 (0.00–0.01) | 0.01 (0.00–0.01) | 0.03 (0.02–0.03) | 0.27 (0.24–0.33) |
|  | **SAC** | 0.02 (0.01–0.03) | 0.05 (0.03–0.07) | 0.06 (0.04–0.08) | 1.35 (1.23–1.53) |
| Blantyre | **Adults** | 0.01 (0.01–0.02) | 0.54 (0.32–0.68) | 0.55 (0.32–0.73) | 2.63 (1.95–3.19) |
|  | **Infant** | 0.00 (0.00–0.00) | 0.00 (0.00–0.00) | 0.00 (0.00–0.00) | 0.06 (0.05–0.08) |
|  | **PSAC** | 0.00 (0.00–0.00) | 0.00 (0.00–0.01) | 0.01 (0.01–0.02) | 0.20 (0.15–0.25) |
|  | **SAC** | 0.01 (0.00–0.01) | 0.02 (0.01–0.03) | 0.03 (0.01–0.04) | 1.09 (0.88–1.27) |
| Blantyre City | **Adults** | 0.02 (0.01–0.03) | 0.85 (0.51–1.15) | 0.88 (0.52–1.14) | 4.16 (3.30–5.08) |
|  | **Infant** | 0.00 (0.00–0.00) | 0.00 (0.00–0.00) | 0.00 (0.00–0.00) | 0.09 (0.07–0.13) |
|  | **PSAC** | 0.00 (0.00–0.01) | 0.01 (0.00–0.01) | 0.02 (0.01–0.03) | 0.31 (0.25–0.39) |
|  | **SAC** | 0.01 (0.00–0.01) | 0.02 (0.01–0.04) | 0.04 (0.02–0.06) | 1.72 (1.48–2.10) |
| Chikwawa | **Adults** | 0.43 (0.37–0.51) | 8.31 (7.90–9.04) | 8.59 (8.19–9.42) | 13.80 (13.22–14.63) |
|  | **Infant** | 0.04 (0.04–0.05) | 0.09 (0.08–0.11) | 0.10 (0.10–0.13) | 0.57 (0.54–0.63) |
|  | **PSAC** | 0.08 (0.07–0.10) | 0.15 (0.14–0.18) | 0.37 (0.35–0.44) | 1.11 (1.05–1.21) |
|  | **SAC** | 0.31 (0.28–0.39) | 0.67 (0.61–0.80) | 0.85 (0.79–1.02) | 4.01 (3.81–4.35) |
| Chiradzulu | **Adults** | 0.00 (0.00–0.00) | 0.09 (0.05–0.15) | 0.11 (0.06–0.19) | 2.13 (1.74–2.83) |
|  | **Infant** | 0.00 (0.00–0.00) | 0.00 (0.00–0.00) | 0.00 (0.00–0.00) | 0.06 (0.04–0.08) |
|  | **PSAC** | 0.00 (0.00–0.00) | 0.00 (0.00–0.00) | 0.01 (0.00–0.01) | 0.17 (0.14–0.23) |
|  | **SAC** | 0.01 (0.00–0.01) | 0.01 (0.01–0.02) | 0.01 (0.01–0.02) | 0.95 (0.82–1.20) |
| Chitipa | **Adults** | 0.01 (0.00–0.01) | 0.25 (0.14–0.38) | 0.26 (0.14–0.40) | 1.28 (0.92–1.60) |
|  | **Infant** | 0.00 (0.00–0.00) | 0.00 (0.00–0.00) | 0.00 (0.00–0.00) | 0.03 (0.02–0.04) |
|  | **PSAC** | 0.00 (0.00–0.00) | 0.00 (0.00–0.00) | 0.01 (0.00–0.01) | 0.10 (0.07–0.12) |
|  | **SAC** | 0.00 (0.00–0.01) | 0.01 (0.00–0.01) | 0.01 (0.01–0.02) | 0.55 (0.44–0.65) |
| Dedza | **Adults** | 0.48 (0.40–0.54) | 10.69 (9.79–11.18) | 10.97 (10.25–11.50) | 18.99 (18.25–19.68) |
|  | **Infant** | 0.05 (0.04–0.05) | 0.10 (0.09–0.12) | 0.12 (0.09–0.13) | 0.78 (0.70–0.86) |
|  | **PSAC** | 0.09 (0.08–0.10) | 0.18 (0.16–0.19) | 0.46 (0.39–0.50) | 1.56 (1.45–1.67) |
|  | **SAC** | 0.35 (0.28–0.42) | 0.82 (0.74–0.91) | 1.02 (0.88–1.12) | 5.64 (5.28–5.95) |
| Dowa | **Adults** | 0.04 (0.03–0.05) | 1.29 (1.02–1.82) | 1.36 (1.04–1.93) | 5.81 (5.08–6.51) |
|  | **Infant** | 0.00 (0.00–0.00) | 0.01 (0.00–0.01) | 0.01 (0.00–0.01) | 0.15 (0.12–0.20) |
|  | **PSAC** | 0.01 (0.00–0.01) | 0.01 (0.01–0.02) | 0.03 (0.02–0.05) | 0.44 (0.38–0.53) |
|  | **SAC** | 0.03 (0.02–0.05) | 0.06 (0.04–0.09) | 0.08 (0.06–0.12) | 2.25 (2.04–2.51) |
| Karonga | **Adults** | 0.01 (0.01–0.02) | 0.48 (0.37–0.60) | 0.51 (0.37–0.63) | 2.69 (2.32–3.20) |
|  | **Infant** | 0.00 (0.00–0.00) | 0.00 (0.00–0.00) | 0.00 (0.00–0.00) | 0.08 (0.06–0.10) |
|  | **PSAC** | 0.00 (0.00–0.01) | 0.00 (0.00–0.01) | 0.02 (0.01–0.02) | 0.22 (0.18–0.27) |
|  | **SAC** | 0.01 (0.01–0.01) | 0.02 (0.02–0.03) | 0.03 (0.03–0.04) | 1.08 (0.97–1.22) |
| Kasungu | **Adults** | 0.53 (0.46–0.58) | 11.73 (10.96–12.36) | 11.99 (11.27–12.73) | 19.41 (18.30–20.25) |
|  | **Infant** | 0.05 (0.04–0.06) | 0.11 (0.09–0.12) | 0.12 (0.10–0.14) | 0.78 (0.71–0.81) |
|  | **PSAC** | 0.09 (0.08–0.11) | 0.18 (0.16–0.21) | 0.47 (0.41–0.52) | 1.55 (1.46–1.64) |
|  | **SAC** | 0.36 (0.31–0.41) | 0.86 (0.76–0.96) | 1.06 (0.92–1.19) | 5.62 (5.40–5.91) |
| Likoma | **Adults** | 0.02 (0.01–0.02) | 0.26 (0.24–0.27) | 0.26 (0.25–0.28) | 0.36 (0.35–0.38) |
|  | **Infant** | 0.00 (0.00–0.00) | 0.00 (0.00–0.00) | 0.00 (0.00–0.00) | 0.01 (0.01–0.02) |
|  | **PSAC** | 0.00 (0.00–0.00) | 0.00 (0.00–0.01) | 0.01 (0.01–0.01) | 0.03 (0.03–0.03) |
|  | **SAC** | 0.01 (0.01–0.01) | 0.02 (0.02–0.02) | 0.02 (0.02–0.03) | 0.10 (0.09–0.11) |
| Lilongwe | **Adults** | 0.08 (0.06–0.10) | 2.91 (2.26–3.71) | 3.04 (2.47–3.97) | 12.90 (11.01–15.77) |
|  | **Infant** | 0.00 (0.00–0.01) | 0.01 (0.01–0.02) | 0.01 (0.01–0.02) | 0.34 (0.26–0.44) |
|  | **PSAC** | 0.02 (0.01–0.02) | 0.03 (0.02–0.04) | 0.09 (0.06–0.12) | 0.99 (0.80–1.20) |
|  | **SAC** | 0.06 (0.03–0.09) | 0.14 (0.08–0.20) | 0.18 (0.12–0.25) | 4.98 (4.34–5.88) |
| Lilongwe City | **Adults** | 0.04 (0.02–0.06) | 1.37 (0.90–2.06) | 1.41 (0.89–2.08) | 5.81 (4.72–7.30) |
|  | **Infant** | 0.00 (0.00–0.00) | 0.00 (0.00–0.01) | 0.01 (0.00–0.01) | 0.13 (0.08–0.17) |
|  | **PSAC** | 0.01 (0.00–0.01) | 0.01 (0.01–0.02) | 0.03 (0.02–0.06) | 0.42 (0.31–0.53) |
|  | **SAC** | 0.02 (0.02–0.04) | 0.05 (0.03–0.09) | 0.07 (0.05–0.11) | 2.31 (1.92–2.77) |
| Machinga | **Adults** | 0.31 (0.22–0.40) | 7.66 (6.47–8.62) | 7.97 (6.71–9.04) | 15.57 (14.54–16.37) |
|  | **Infant** | 0.03 (0.03–0.05) | 0.07 (0.05–0.08) | 0.08 (0.07–0.10) | 0.64 (0.58–0.72) |
|  | **PSAC** | 0.06 (0.05–0.08) | 0.12 (0.10–0.14) | 0.34 (0.29–0.39) | 1.30 (1.20–1.43) |
|  | **SAC** | 0.25 (0.19–0.30) | 0.58 (0.49–0.69) | 0.75 (0.61–0.87) | 4.73 (4.43–5.07) |
| Mangochi | **Adults** | 0.90 (0.65–1.07) | 17.17 (14.47–18.98) | 17.66 (14.86–19.36) | 27.65 (25.95–29.12) |
|  | **Infant** | 0.09 (0.06–0.12) | 0.17 (0.12–0.21) | 0.21 (0.15–0.25) | 1.13 (1.04–1.29) |
|  | **PSAC** | 0.17 (0.12–0.21) | 0.29 (0.22–0.36) | 0.77 (0.61–0.87) | 2.24 (2.06–2.53) |
|  | **SAC** | 0.61 (0.46–0.76) | 1.32 (1.00–1.57) | 1.74 (1.33–2.01) | 7.95 (7.40–8.81) |
| Mchinji | **Adults** | 0.04 (0.03–0.06) | 1.52 (1.09–1.96) | 1.56 (1.10–1.93) | 4.85 (3.95–5.68) |
|  | **Infant** | 0.00 (0.00–0.00) | 0.01 (0.00–0.01) | 0.01 (0.00–0.01) | 0.13 (0.09–0.16) |
|  | **PSAC** | 0.00 (0.00–0.01) | 0.01 (0.01–0.01) | 0.04 (0.02–0.06) | 0.37 (0.30–0.43) |
|  | **SAC** | 0.02 (0.01–0.04) | 0.07 (0.05–0.10) | 0.08 (0.05–0.12) | 1.81 (1.56–2.00) |
| Mulanje | **Adults** | 0.15 (0.09–0.19) | 4.38 (3.20–5.26) | 4.67 (3.31–5.61) | 13.41 (11.59–14.76) |
|  | **Infant** | 0.02 (0.01–0.03) | 0.04 (0.03–0.05) | 0.04 (0.02–0.06) | 0.56 (0.48–0.63) |
|  | **PSAC** | 0.04 (0.02–0.05) | 0.07 (0.05–0.09) | 0.21 (0.13–0.26) | 1.17 (1.03–1.29) |
|  | **SAC** | 0.16 (0.10–0.22) | 0.34 (0.24–0.44) | 0.46 (0.28–0.59) | 4.33 (3.86–4.68) |
| Mwanza | **Adults** | 0.00 (0.00–0.00) | 0.11 (0.07–0.16) | 0.11 (0.07–0.16) | 0.71 (0.56–0.88) |
|  | **Infant** | 0.00 (0.00–0.00) | 0.00 (0.00–0.00) | 0.00 (0.00–0.00) | 0.02 (0.01–0.02) |
|  | **PSAC** | 0.00 (0.00–0.00) | 0.00 (0.00–0.00) | 0.00 (0.00–0.01) | 0.06 (0.04–0.07) |
|  | **SAC** | 0.00 (0.00–0.00) | 0.00 (0.00–0.01) | 0.01 (0.00–0.01) | 0.31 (0.25–0.36) |
| Mzimba | **Adults** | 0.03 (0.01–0.05) | 1.02 (0.50–1.77) | 1.06 (0.50–1.81) | 5.36 (4.07–6.81) |
|  | **Infant** | 0.00 (0.00–0.00) | 0.00 (0.00–0.01) | 0.00 (0.00–0.01) | 0.14 (0.10–0.18) |
|  | **PSAC** | 0.00 (0.00–0.01) | 0.01 (0.00–0.02) | 0.03 (0.02–0.05) | 0.42 (0.34–0.52) |
|  | **SAC** | 0.01 (0.01–0.02) | 0.04 (0.02–0.06) | 0.05 (0.02–0.09) | 2.25 (1.89–2.57) |
| Mzuzu City | **Adults** | 0.00 (0.00–0.01) | 0.13 (0.07–0.18) | 0.13 (0.08–0.20) | 0.65 (0.52–0.77) |
|  | **Infant** | 0.00 (0.00–0.00) | 0.00 (0.00–0.00) | 0.00 (0.00–0.00) | 0.02 (0.01–0.02) |
|  | **PSAC** | 0.00 (0.00–0.00) | 0.00 (0.00–0.00) | 0.00 (0.00–0.00) | 0.05 (0.04–0.07) |
|  | **SAC** | 0.00 (0.00–0.00) | 0.00 (0.00–0.01) | 0.01 (0.00–0.01) | 0.27 (0.23–0.33) |
| Neno | **Adults** | 0.01 (0.00–0.01) | 0.25 (0.17–0.35) | 0.26 (0.18–0.37) | 1.07 (0.87–1.21) |
|  | **Infant** | 0.00 (0.00–0.00) | 0.00 (0.00–0.00) | 0.00 (0.00–0.00) | 0.03 (0.02–0.04) |
|  | **PSAC** | 0.00 (0.00–0.00) | 0.00 (0.00–0.00) | 0.01 (0.00–0.01) | 0.08 (0.07–0.10) |
|  | **SAC** | 0.01 (0.00–0.01) | 0.01 (0.01–0.02) | 0.02 (0.01–0.02) | 0.41 (0.36–0.47) |
| Nkhata Bay | **Adults** | 0.01 (0.01–0.02) | 0.39 (0.30–0.58) | 0.41 (0.30–0.59) | 2.05 (1.77–2.25) |
|  | **Infant** | 0.00 (0.00–0.00) | 0.00 (0.00–0.00) | 0.00 (0.00–0.00) | 0.05 (0.04–0.06) |
|  | **PSAC** | 0.00 (0.00–0.00) | 0.00 (0.00–0.00) | 0.01 (0.01–0.01) | 0.16 (0.14–0.18) |
|  | **SAC** | 0.01 (0.01–0.01) | 0.02 (0.01–0.02) | 0.02 (0.02–0.03) | 0.82 (0.74–0.88) |
| Nkhotakota | **Adults** | 0.00 (0.00–0.00) | 0.07 (0.03–0.11) | 0.08 (0.04–0.12) | 2.06 (1.68–2.35) |
|  | **Infant** | 0.00 (0.00–0.00) | 0.00 (0.00–0.00) | 0.00 (0.00–0.00) | 0.05 (0.04–0.07) |
|  | **PSAC** | 0.00 (0.00–0.00) | 0.00 (0.00–0.00) | 0.01 (0.00–0.01) | 0.17 (0.13–0.21) |
|  | **SAC** | 0.01 (0.00–0.01) | 0.01 (0.00–0.01) | 0.01 (0.01–0.02) | 0.95 (0.82–1.04) |
| Nsanje | **Adults** | 0.13 (0.10–0.17) | 3.07 (2.75–3.58) | 3.23 (2.91–3.72) | 6.58 (6.23–6.97) |
|  | **Infant** | 0.01 (0.01–0.02) | 0.03 (0.02–0.04) | 0.04 (0.03–0.05) | 0.27 (0.25–0.30) |
|  | **PSAC** | 0.03 (0.02–0.04) | 0.05 (0.04–0.07) | 0.14 (0.11–0.18) | 0.55 (0.51–0.59) |
|  | **SAC** | 0.11 (0.09–0.14) | 0.24 (0.19–0.31) | 0.31 (0.26–0.39) | 2.00 (1.88–2.12) |
| Ntcheu | **Adults** | 0.04 (0.03–0.06) | 1.44 (1.07–2.02) | 1.51 (1.08–2.08) | 5.40 (5.00–5.87) |
|  | **Infant** | 0.00 (0.00–0.00) | 0.01 (0.00–0.01) | 0.01 (0.00–0.01) | 0.14 (0.11–0.17) |
|  | **PSAC** | 0.01 (0.00–0.01) | 0.01 (0.01–0.02) | 0.04 (0.03–0.06) | 0.40 (0.36–0.46) |
|  | **SAC** | 0.03 (0.02–0.04) | 0.06 (0.05–0.09) | 0.09 (0.06–0.13) | 2.01 (1.84–2.18) |
| Ntchisi | **Adults** | 0.02 (0.01–0.02) | 0.64 (0.52–0.76) | 0.66 (0.53–0.78) | 2.07 (1.80–2.37) |
|  | **Infant** | 0.00 (0.00–0.00) | 0.00 (0.00–0.00) | 0.00 (0.00–0.00) | 0.05 (0.03–0.06) |
|  | **PSAC** | 0.00 (0.00–0.00) | 0.00 (0.00–0.01) | 0.01 (0.01–0.02) | 0.15 (0.11–0.18) |
|  | **SAC** | 0.00 (0.00–0.01) | 0.02 (0.01–0.02) | 0.03 (0.02–0.03) | 0.81 (0.67–0.91) |
| Phalombe | **Adults** | 0.31 (0.29–0.34) | 6.80 (6.41–7.14) | 6.92 (6.58–7.29) | 10.38 (10.13–10.70) |
|  | **Infant** | 0.02 (0.02–0.03) | 0.06 (0.05–0.07) | 0.07 (0.06–0.08) | 0.41 (0.38–0.43) |
|  | **PSAC** | 0.05 (0.04–0.05) | 0.10 (0.09–0.11) | 0.26 (0.25–0.29) | 0.83 (0.77–0.86) |
|  | **SAC** | 0.17 (0.15–0.20) | 0.46 (0.41–0.52) | 0.58 (0.54–0.66) | 2.98 (2.85–3.07) |
| Rumphi | **Adults** | 0.01 (0.01–0.01) | 0.28 (0.21–0.42) | 0.30 (0.23–0.43) | 1.59 (1.37–1.90) |
|  | **Infant** | 0.00 (0.00–0.00) | 0.00 (0.00–0.00) | 0.00 (0.00–0.00) | 0.04 (0.03–0.05) |
|  | **PSAC** | 0.00 (0.00–0.00) | 0.00 (0.00–0.00) | 0.01 (0.01–0.01) | 0.12 (0.10–0.14) |
|  | **SAC** | 0.01 (0.01–0.01) | 0.01 (0.01–0.02) | 0.02 (0.01–0.03) | 0.63 (0.56–0.71) |
| Salima | **Adults** | 0.00 (0.00–0.00) | 0.05 (0.01–0.10) | 0.06 (0.02–0.09) | 2.34 (1.83–2.91) |
|  | **Infant** | 0.00 (0.00–0.00) | 0.00 (0.00–0.00) | 0.00 (0.00–0.00) | 0.06 (0.04–0.08) |
|  | **PSAC** | 0.00 (0.00–0.00) | 0.00 (0.00–0.00) | 0.01 (0.00–0.01) | 0.20 (0.15–0.26) |
|  | **SAC** | 0.01 (0.00–0.01) | 0.01 (0.00–0.01) | 0.01 (0.01–0.01) | 1.10 (0.90–1.31) |
| Thyolo | **Adults** | 0.01 (0.01–0.02) | 0.50 (0.39–0.74) | 0.58 (0.43–0.88) | 4.55 (4.00–5.22) |
|  | **Infant** | 0.00 (0.00–0.00) | 0.00 (0.00–0.00) | 0.00 (0.00–0.01) | 0.12 (0.09–0.14) |
|  | **PSAC** | 0.00 (0.00–0.01) | 0.01 (0.00–0.01) | 0.02 (0.01–0.03) | 0.37 (0.30–0.42) |
|  | **SAC** | 0.02 (0.02–0.03) | 0.03 (0.02–0.04) | 0.05 (0.03–0.07) | 1.92 (1.72–2.12) |
| Zomba | **Adults** | 0.44 (0.36–0.50) | 9.68 (8.79–10.56) | 10.03 (9.03–10.88) | 17.41 (16.12–18.30) |
|  | **Infant** | 0.05 (0.04–0.05) | 0.09 (0.07–0.11) | 0.11 (0.09–0.12) | 0.73 (0.65–0.79) |
|  | **PSAC** | 0.09 (0.07–0.10) | 0.16 (0.13–0.19) | 0.42 (0.37–0.47) | 1.45 (1.33–1.55) |
|  | **SAC** | 0.33 (0.23–0.38) | 0.73 (0.60–0.84) | 0.95 (0.80–1.03) | 5.22 (4.85–5.44) |
| Zomba City | **Adults** | 0.06 (0.06–0.07) | 1.35 (1.27–1.41) | 1.37 (1.29–1.43) | 2.33 (2.19–2.42) |
|  | **Infant** | 0.01 (0.00–0.01) | 0.01 (0.01–0.01) | 0.01 (0.01–0.02) | 0.09 (0.08–0.10) |
|  | **PSAC** | 0.01 (0.01–0.01) | 0.02 (0.02–0.02) | 0.05 (0.04–0.06) | 0.18 (0.17–0.19) |
|  | **SAC** | 0.04 (0.03–0.04) | 0.09 (0.08–0.10) | 0.12 (0.10–0.13) | 0.67 (0.62–0.71) |

*Values represent the mean and 95% confidence intervals across ten runs for each draw.*

Table 6. Mean undiscounted costs, 2024-2050 in USD (millions).

|  | Total costs of MDA to treat target population (millions) | | | | | |
| --- | --- | --- | --- | --- | --- | --- |
|  | MDA All | | MSA PSAC+SAC | | MDA SAC | |
| District  *Population size in 2024 (millions)* | Consumables | Financial costs | Consumables | Financial costs | Consumables | Financial costs |
| Balaka  *0.54* | 0.10 (0.10–0.10) | 0.78 (0.77–0.80) | 0.22 (0.22–0.24) | 2.92 (2.84–3.06) | 0.17 (0.16–0.17) | 2.20 (2.14–2.27) |
| Blantyre  *0.53* | 0.06 (0.06–0.06) | 0.50 (0.48–0.52) | 0.22 (0.21–0.23) | 2.85 (2.67–3.02) | 0.17 (0.16–0.17) | 2.15 (2.04–2.26) |
| Blantyre City  *0.94* | 0.11 (0.11–0.12) | 0.90 (0.88–0.92) | 0.39 (0.37–0.40) | 5.05 (4.75–5.24) | 0.30 (0.28–0.31) | 3.84 (3.60–4.03) |
| Chikwawa  *0.68* | 0.35 (0.34–0.36) | 2.79 (2.72–2.90) | 0.28 (0.27–0.30) | 3.65 (3.50–3.90) | 0.21 (0.20–0.22) | 2.75 (2.63–2.87) |
| Chiradzulu  *0.44* | 0.05 (0.05–0.06) | 0.42 (0.41–0.44) | 0.01 (0.01–0.01) | 0.14 (0.14–0.16) | 0.01 (0.01–0.01) | 0.16 (0.16–0.18) |
| Chitipa  *0.28* | 0.03 (0.03–0.03) | 0.27 (0.25–0.27) | 0.12 (0.11–0.12) | 1.52 (1.42–1.59) | 0.09 (0.08–0.09) | 1.15 (1.07–1.21) |
| Dedza  *0.99* | 0.44 (0.42–0.45) | 3.50 (3.38–3.59) | 0.41 (0.39–0.43) | 5.34 (5.11–5.58) | 0.31 (0.29–0.33) | 4.05 (3.82–4.27) |
| Dowa  *0.93* | 0.17 (0.16–0.17) | 1.35 (1.30–1.40) | 0.39 (0.37–0.41) | 5.05 (4.80–5.27) | 0.29 (0.27–0.31) | 3.80 (3.52–4.08) |
| Karonga  *0.44* | 0.08 (0.08–0.08) | 0.63 (0.61–0.65) | 0.18 (0.18–0.19) | 2.40 (2.30–2.53) | 0.14 (0.13–0.15) | 1.82 (1.72–1.92) |
| Kasungu  *0.99* | 0.44 (0.42–0.45) | 3.53 (3.40–3.63) | 0.41 (0.40–0.42) | 5.37 (5.19–5.50) | 0.31 (0.30–0.32) | 4.04 (3.89–4.18) |
| Likoma  *0.02* | 0.01 (0.01–0.01) | 0.07 (0.07–0.07) | 0.01 (0.01–0.01) | 0.09 (0.09–0.10) | 0.01 (0.01–0.01) | 0.07 (0.07–0.07) |
| Lilongwe  *1.98* | 0.36 (0.35–0.36) | 2.86 (2.80–2.91) | 0.83 (0.80–0.86) | 10.73 (10.37–11.14) | 0.62 (0.59–0.65) | 8.09 (7.73–8.46) |
| Lilongwe City  *1.16* | 0.21 (0.20–0.21) | 1.67 (1.63–1.71) | 0.48 (0.46–0.51) | 6.28 (5.96–6.57) | 0.36 (0.35–0.38) | 4.71 (4.55–4.92) |
| Machinga  *0.84* | 0.32 (0.31–0.33) | 2.54 (2.46–2.63) | 0.35 (0.34–0.37) | 4.56 (4.40–4.80) | 0.27 (0.25–0.28) | 3.45 (3.30–3.64) |
| Mangochi  *1.37* | 0.70 (0.67–0.74) | 5.62 (5.36–5.91) | 0.57 (0.53–0.62) | 7.43 (6.90–8.11) | 0.43 (0.40–0.46) | 5.58 (5.26–6.02) |
| Mchinji  *0.73* | 0.13 (0.13–0.14) | 1.06 (1.02–1.09) | 0.31 (0.28–0.32) | 3.97 (3.62–4.15) | 0.23 (0.22–0.25) | 2.99 (2.80–3.19) |
| Mulanje  *0.81* | 0.30 (0.29–0.32) | 2.44 (2.36–2.55) | 0.34 (0.32–0.36) | 4.36 (4.20–4.62) | 0.25 (0.24–0.27) | 3.28 (3.11–3.53) |
| Mwanza  *0.15* | 0.02 (0.02–0.02) | 0.15 (0.14–0.15) | 0.06 (0.06–0.07) | 0.84 (0.79–0.89) | 0.05 (0.05–0.05) | 0.63 (0.60–0.67) |
| Mzimba  *1.09* | 0.13 (0.13–0.13) | 1.04 (1.01–1.07) | 0.45 (0.42–0.47) | 5.88 (5.49–6.11) | 0.34 (0.32–0.36) | 4.46 (4.18–4.66) |
| Mzuzu City  *0.25* | 0.03 (0.03–0.03) | 0.24 (0.23–0.25) | 0.11 (0.10–0.11) | 1.37 (1.30–1.43) | 0.08 (0.08–0.08) | 1.04 (0.99–1.08) |
| Neno  *0.16* | 0.03 (0.03–0.03) | 0.23 (0.23–0.24) | 0.07 (0.06–0.07) | 0.88 (0.82–0.93) | 0.05 (0.05–0.05) | 0.66 (0.62–0.71) |
| Nkhata Bay  *0.35* | 0.06 (0.06–0.07) | 0.51 (0.49–0.53) | 0.15 (0.14–0.15) | 1.90 (1.80–2.01) | 0.11 (0.11–0.12) | 1.45 (1.37–1.53) |
| Nkhotakota  *0.46* | 0.00 (0.00–0.00) | 0.00 (0.00–0.00) | 0.00 (0.00–0.00) | 0.00 (0.00–0.00) | 0.00 (0.00–0.00) | 0.00 (0.00–0.00) |
| Nsanje  *0.36* | 0.16 (0.15–0.16) | 1.27 (1.24–1.31) | 0.15 (0.14–0.16) | 1.94 (1.86–2.02) | 0.11 (0.11–0.12) | 1.47 (1.41–1.53) |
| Ntcheu  *0.80* | 0.14 (0.14–0.15) | 1.16 (1.12–1.20) | 0.33 (0.32–0.35) | 4.34 (4.11–4.57) | 0.25 (0.24–0.27) | 3.28 (3.11–3.45) |
| Ntchisi  *0.38* | 0.07 (0.07–0.07) | 0.54 (0.52–0.57) | 0.16 (0.15–0.17) | 2.07 (1.94–2.15) | 0.12 (0.11–0.12) | 1.55 (1.46–1.62) |
| Phalombe  *0.52* | 0.23 (0.22–0.24) | 1.84 (1.77–1.91) | 0.22 (0.21–0.23) | 2.83 (2.74–2.98) | 0.16 (0.16–0.17) | 2.12 (2.05–2.20) |
| Rumphi  *0.27* | 0.05 (0.05–0.05) | 0.39 (0.38–0.40) | 0.11 (0.11–0.12) | 1.47 (1.40–1.54) | 0.09 (0.08–0.09) | 1.11 (1.07–1.14) |
| Salima  *0.56* | 0.00 (0.00–0.00) | 0.00 (0.00–0.00) | 0.00 (0.00–0.00) | 0.00 (0.00–0.00) | 0.00 (0.00–0.00) | 0.00 (0.00–0.00) |
| Thyolo  *0.84* | 0.15 (0.15–0.15) | 1.21 (1.17–1.24) | 0.26 (0.25–0.27) | 3.41 (3.27–3.56) | 0.26 (0.25–0.27) | 3.41 (3.29–3.54) |
| Zomba  *0.89* | 0.40 (0.38–0.41) | 3.17 (3.08–3.31) | 0.37 (0.35–0.39) | 4.83 (4.58–5.07) | 0.28 (0.27–0.30) | 3.65 (3.45–3.84) |
| Zomba City  *0.13* | 0.06 (0.05–0.06) | 0.45 (0.42–0.47) | 0.05 (0.05–0.06) | 0.69 (0.65–0.73) | 0.04 (0.04–0.04) | 0.52 (0.49–0.55) |

*Financial costs here relate to implementation costs only and exclude consumables. In some districts, costs for “MDA All” are lower than for “PSAC+SAC” because the district reaches elimination earlier and MDA ceases.*

Table 7. District-level net health benefit relative to no MDA (2024 - 2050) under alternative WASH and MDA strategies.

| **District** | **Continue WASH** | | | **Pause WASH** | | | **Scale-up WASH** | | |
| --- | --- | --- | --- | --- | --- | --- | --- | --- | --- |
|  | **MDA SAC** | **MDA PSAC+SAC** | **MDA All** | **MDA SAC** | **MDA PSAC+SAC** | **MDA All** | **MDA SAC** | **MDA PSAC+SAC** | **MDA All** |
| Balaka | 49002.27 (32038.15–65966.40) | 40207.13 (23373.54–57040.72) | 65937.12 (49067.52–82806.72) | 56685.37 (34130.81–79239.93) | 47878.45 (25293.60–70463.29) | 73785.78 (51083.01–96488.55) | 24365.26 (9947.55–38782.96) | 15288.43 (825.86–29751.01) | 41404.96 (26912.50–55897.42) |
| Blantyre | 8561.84 (-1337.61–18461.28) | -34.60 (-9597.71–9528.50) | 28457.67 (18254.18–38661.15) | 14524.60 (-1941.60–30990.81) | 6479.49 (-9992.64–22951.62) | 34649.77 (18021.70–51277.83) | -3564.71 (-12104.59–4975.17) | -12351.36 (-20694.43–-4008.29) | 16274.34 (7393.53–25155.15) |
| Blantyre City | 2452.24 (-15452.50–20356.97) | -12365.23 (-30185.68–5455.23) | 37908.48 (20044.07–55772.88) | -6480.17 (-31661.66–18701.31) | -22012.77 (-47181.03–3155.49) | 28927.34 (3395.60–54459.09) | -13517.53 (-25697.23–-1337.82) | -29418.42 (-41525.24–-17311.59) | 21720.13 (9333.43–34106.84) |
| Chikwawa | 1739985.09 (1635953.32–1844016.86) | 1737064.82 (1632871.23–1841258.41) | 1762363.16 (1656888.63–1867837.70) | 1732300.66 (1635006.06–1829595.27) | 1729041.70 (1630811.37–1827272.03) | 1754315.03 (1654452.26–1854177.80) | 1611473.71 (1525696.80–1697250.62) | 1608728.13 (1523209.17–1694247.10) | 1633565.39 (1547645.63–1719485.15) |
| Chiradzulu | 27954.00 (14172.33–41735.67) | 28199.28 (14417.21–41981.34) | 24581.11 (10824.82–38337.40) | 42542.91 (27962.03–57123.79) | 42785.21 (28202.55–57367.87) | 39163.91 (24622.77–53705.05) | 19519.04 (7647.61–31390.47) | 19752.03 (7885.58–31618.47) | 16131.79 (4307.04–27956.55) |
| Chitipa | 1009.99 (-6197.63–8217.61) | -3587.85 (-10718.71–3543.01) | 11643.65 (4330.69–18956.61) | 5023.87 (-2214.18–12261.91) | 486.09 (-6835.47–7807.65) | 15684.64 (8404.73–22964.56) | -4197.06 (-8380.26–-13.85) | -8798.33 (-12873.38–-4723.27) | 6375.89 (2206.81–10544.97) |
| Dedza | 2113892.52 (1932675.40–2295109.65) | 2101834.94 (1920104.58–2283565.30) | 2132619.55 (1950555.13–2314683.97) | 2054981.14 (1919501.99–2190460.28) | 2042264.86 (1906579.89–2177949.83) | 2073290.11 (1937004.82–2209575.39) | 1942529.47 (1803587.01–2081471.94) | 1931110.39 (1791707.79–2070512.99) | 1961197.85 (1821646.52–2100749.19) |
| Dowa | 59429.04 (29987.21–88870.87) | 44101.40 (14377.05–73825.75) | 88643.82 (58617.18–118670.46) | 64068.60 (38633.48–89503.72) | 48910.11 (23751.71–74068.51) | 93313.44 (67246.46–119380.41) | 38055.29 (8709.80–67400.79) | 22453.20 (-6834.46–51740.86) | 67552.09 (37708.09–97396.10) |
| Karonga | 41552.58 (19081.22–64023.93) | 34456.06 (11835.64–57076.47) | 55703.83 (33256.59–78151.06) | 31385.24 (17351.34–45419.14) | 24265.39 (10284.67–38246.11) | 45485.76 (31372.79–59598.74) | 23280.91 (7415.32–39146.49) | 15941.83 (154.94–31728.72) | 37381.53 (21511.72–53251.34) |
| Kasungu | 2126853.96 (1976297.03–2277410.90) | 2115448.45 (1964397.02–2266499.87) | 2148068.16 (1996509.86–2299626.46) | 2080585.99 (1915285.41–2245886.57) | 2069335.65 (1902976.98–2235694.32) | 2102085.27 (1934970.79–2269199.74) | 1905033.12 (1752545.72–2057520.51) | 1894183.56 (1740997.97–2047369.14) | 1926488.79 (1772689.18–2080288.41) |
| Likoma | 43315.94 (40688.48–45943.39) | 43392.16 (40743.43–46040.89) | 44746.88 (41991.64–47502.13) | 43065.86 (40307.17–45824.55) | 43145.24 (40334.56–45955.92) | 44515.65 (41548.01–47483.29) | 38810.06 (35674.63–41945.50) | 38869.01 (35696.58–42041.45) | 40232.57 (36937.33–43527.81) |
| Lilongwe | 212163.53 (121698.26–302628.81) | 179906.17 (89445.66–270366.67) | 274656.59 (184160.61–365152.57) | 149019.06 (83238.38–214799.73) | 117791.95 (52172.13–183411.77) | 212200.49 (145712.54–278688.45) | 125749.71 (50730.30–200769.12) | 93916.88 (19390.22–168443.55) | 188635.50 (112982.19–264288.80) |
| Lilongwe City | 10702.83 (-12514.99–33920.65) | -8450.20 (-31585.09–14684.69) | 46994.74 (22950.21–71039.27) | 15385.98 (-18239.67–49011.63) | -3542.75 (-36775.18–29689.69) | 51892.15 (17826.52–85957.78) | -15616.25 (-34423.72–3191.21) | -34832.35 (-53429.87–-16234.83) | 20631.69 (1521.24–39742.13) |
| Machinga | 1636297.76 (1525427.25–1747168.27) | 1624505.13 (1513217.50–1735792.76) | 1651053.50 (1538305.98–1763801.03) | 1642103.51 (1536554.83–1747652.19) | 1630007.36 (1523819.95–1736194.78) | 1656787.88 (1549074.78–1764500.99) | 1488533.56 (1327780.61–1649286.52) | 1476470.68 (1315195.46–1637745.90) | 1503126.35 (1340592.89–1665659.82) |
| Mangochi | 3275168.03 (2996546.98–3553789.07) | 3269805.15 (2990260.35–3549349.95) | 3327832.61 (3042091.04–3613574.17) | 3362669.42 (3053326.23–3672012.61) | 3356916.35 (3044507.26–3669325.43) | 3414149.29 (3095604.20–3732694.39) | 2967201.80 (2695321.34–3239082.25) | 2961780.54 (2687696.47–3235864.62) | 3019542.76 (2739979.09–3299106.42) |
| Mchinji | 61326.28 (33051.61–89600.95) | 49303.82 (21315.08–77292.55) | 84385.06 (55626.41–113143.72) | 57795.62 (38575.90–77015.34) | 45767.62 (26757.35–64777.89) | 80993.56 (61524.78–100462.34) | 38789.22 (14822.97–62755.46) | 26936.69 (3257.80–50615.59) | 61999.53 (37763.73–86235.33) |
| Mulanje | 1300706.81 (1150244.95–1451168.67) | 1287707.12 (1137253.24–1438161.00) | 1310083.13 (1158961.95–1461204.30) | 1277920.11 (1110989.19–1444851.03) | 1265033.36 (1098161.18–1431905.55) | 1287371.77 (1119904.76–1454838.77) | 1122803.97 (964742.09–1280865.86) | 1110091.14 (951749.72–1268432.55) | 1132377.97 (973592.25–1291163.69) |
| Mwanza | 1119.19 (-2960.28–5198.67) | -1399.43 (-5502.95–2704.09) | 7012.19 (2922.46–11101.93) | 947.33 (-1438.27–3332.92) | -1100.90 (-3503.30–1301.50) | 6860.34 (4417.04–9303.64) | -1308.45 (-5229.77–2612.87) | -3742.94 (-7730.11–244.23) | 4611.67 (675.35–8548.00) |
| Mzimba | 36276.88 (13589.59–58964.17) | 18838.00 (-3790.90–41466.89) | 77530.76 (54580.61–100480.92) | 19294.18 (-432.76–39021.12) | 2613.02 (-16679.01–21905.05) | 60816.32 (40872.11–80760.52) | 4836.30 (-17531.39–27203.99) | -13220.06 (-35352.69–8912.58) | 45987.08 (24081.36–67892.80) |
| Mzuzu City | -4234.30 (-7699.15–-769.44) | -8275.09 (-11712.69–-4837.48) | 5397.64 (1936.17–8859.11) | -2564.38 (-6204.68–1075.93) | -6667.46 (-10254.14–-3080.79) | 7100.19 (3399.71–10800.67) | -6869.97 (-9264.75–-4475.19) | -10944.26 (-13423.98–-8464.54) | 2752.28 (273.70–5230.86) |
| Neno | 16348.91 (11527.94–21169.88) | 13647.90 (8782.95–18512.85) | 21461.17 (16562.95–26359.39) | 17296.54 (10513.36–24079.73) | 14679.63 (7941.73–21417.52) | 22404.01 (15522.52–29285.49) | 10696.18 (6532.77–14859.59) | 8066.49 (3897.38–12235.59) | 15831.23 (11625.52–20036.95) |
| Nkhata Bay | 26483.11 (18337.21–34629.02) | 20934.21 (12906.68–28961.73) | 37685.24 (29445.59–45924.88) | 28233.38 (18827.90–37638.86) | 22649.49 (13434.31–31864.68) | 39393.94 (29987.90–48799.99) | 8064.18 (1165.17–14963.18) | 2509.11 (-4287.79–9306.01) | 19279.45 (12252.04–26306.86) |
| Nkhotakota | 23626.38 (12888.82–34363.94) | 23626.38 (12888.82–34363.94) | 23626.38 (12888.82–34363.94) | 29751.38 (19409.67–40093.08) | 29751.38 (19409.67–40093.08) | 29751.38 (19409.67–40093.08) | 15284.93 (11202.50–19367.36) | 15284.93 (11202.50–19367.36) | 15284.93 (11202.50–19367.36) |
| Nsanje | 722879.86 (640481.09–805278.63) | 718253.77 (635540.96–800966.58) | 727029.29 (643764.15–810294.42) | 729305.38 (637674.38–820936.37) | 724490.05 (632424.87–816555.23) | 733224.60 (640599.46–825849.75) | 644606.11 (567594.42–721617.80) | 639809.59 (562375.86–717243.32) | 648750.73 (570828.31–726673.16) |
| Ntcheu | 84381.29 (48974.69–119787.89) | 71454.39 (35948.83–106959.95) | 109751.77 (74327.82–145175.71) | 61782.31 (37982.97–85581.65) | 48992.24 (25077.85–72906.62) | 87215.81 (63131.80–111299.83) | 54995.87 (26967.88–83023.87) | 41548.86 (13695.56–69402.17) | 80422.31 (52191.50–108653.12) |
| Ntchisi | 7428.23 (-1951.30–16807.76) | 1135.77 (-8102.91–10374.45) | 19478.35 (9900.13–29056.57) | 10340.61 (2277.90–18403.31) | 3992.29 (-4152.02–12136.61) | 22384.84 (14256.39–30513.30) | -17.14 (-9760.66–9726.37) | -6013.49 (-15783.64–3756.66) | 12003.40 (2109.91–21896.88) |
| Phalombe | 1117346.77 (1062797.37–1171896.17) | 1111292.85 (1056106.45–1166479.26) | 1128934.87 (1072798.32–1185071.41) | 1162603.15 (1100701.15–1224505.16) | 1156954.40 (1094847.72–1219061.09) | 1174480.09 (1111907.11–1237053.07) | 960144.65 (897783.89–1022505.40) | 954151.31 (891317.63–1016984.99) | 971781.71 (907890.49–1035672.92) |
| Rumphi | 15837.74 (9527.00–22148.49) | 11436.35 (5141.63–17731.07) | 24428.04 (18193.06–30663.02) | 16474.60 (8886.10–24063.10) | 12068.11 (4513.86–19622.36) | 25101.66 (17509.08–32694.24) | 3981.20 (-2441.44–10403.84) | -447.70 (-6945.43–6050.04) | 12559.69 (6122.99–18996.38) |
| Salima | 36515.80 (21035.81–51995.79) | 36515.80 (21035.81–51995.79) | 36515.80 (21035.81–51995.79) | 24350.74 (18530.01–30171.47) | 24350.74 (18530.01–30171.47) | 24350.74 (18530.01–30171.47) | 20613.84 (10365.92–30861.77) | 20613.84 (10365.92–30861.77) | 20613.84 (10365.92–30861.77) |
| Thyolo | 38323.28 (16035.17–60611.38) | 38332.73 (16008.38–60657.08) | 64622.26 (42596.81–86647.71) | 51489.46 (19991.80–82987.12) | 44913.25 (13020.01–76806.50) | 77814.70 (46340.36–109289.04) | 28866.98 (-5256.91–62990.87) | 19998.62 (-14345.87–54343.11) | 55109.17 (21186.05–89032.28) |
| Zomba | 1972730.15 (1818631.80–2126828.49) | 1962423.46 (1807605.85–2117241.06) | 1988284.02 (1831514.96–2145053.07) | 1997398.43 (1847256.66–2147540.19) | 1987607.41 (1836672.47–2138542.35) | 2013359.78 (1860991.10–2165728.47) | 1767803.24 (1579347.80–1956258.68) | 1757549.50 (1567841.40–1947257.61) | 1783703.02 (1592458.65–1974947.39) |
| Zomba City | 219705.74 (204171.22–235240.26) | 218044.39 (202514.75–233574.04) | 221695.93 (206169.01–237222.84) | 222436.92 (204951.99–239921.86) | 220771.43 (203291.90–238250.97) | 224445.08 (206913.44–241976.72) | 214398.21 (198123.94–230672.48) | 212710.36 (196458.17–228962.55) | 216348.25 (200059.43–232637.06) |

Values are presented as mean estimates with 95% uncertainty intervals. Net health benefit was calculated using a willingness-to-pay threshold of 88 USD per disability-adjusted life year (DALY) averted. Negative values indicate that incremental costs exceed the value of health gains.

### H. Sensitivity analysis

A series of sensitivity analyses were conducted to assess the robustness of cost-effectiveness results to alternative cost assumptions, WASH assumptions, discounting choices, disability weights and decision thresholds. All analyses were performed using the same model structure and epidemiological assumptions as in the main results, with only the specified economic or lifestyle inputs varied. For each scenario, we additionally calculated (i) the maximum allowable costs (Table 1), (ii) the district-weighted ICER and (iii) the proportion of districts in which the ICER fell below the Malawi cost-effectiveness threshold of USD 88 per DALY (Table 2). Additionally, for the opportunity cost threshold analysis, Figure 1 presents the optimal district-level strategy across the full threshold range (from USD 1 - 11).

##### Scenario scale-up of WASH access

To examine sensitivity of schistosomiasis outcomes to evolving WASH conditions, we applied externally defined WASH scale-up trajectories by modifying the transition rates described above. Three WASH scenarios were evaluated:

1. Pause WASH (no further expansion):
   - All transition rates were set to zero from 2023 onwards
   - District-level WASH coverage remains fixed at 2023 levels
2. Continue WASH (baseline secular trend):
   - The model’s default secular improvement rates (the baseline three-monthly transition probabilities) are retained without modification
   - WASH coverage therefore evolves gradually over time, driven by the endogenous lifestyle module dynamics
3. Scale-up WASH (accelerated improvement):
   - WASH access is immediately set to universal coverage at the start of the scenario
   - No subsequent decline in access is permitted

##### Alternative cost specifications

In the primary economic analysis, incremental cost-effectiveness ratios (ICERs) were calculated using consumables-only costs, reflecting the variable component of expenditure that scales most directly with the number of individuals treated. To explore the influence of broader programme costs on cost-effectiveness, we conducted two additional scenario analyses: (i) inclusion of partial programme costs, and (ii) inclusion of full economic costs (consumables, programme delivery costs, and human resources).

###### Programme delivery and HR cost components

Programme delivery costs represent the economic cost of planning, community sensitisation, training, management, supervision, and drug distribution. These costs include both routine financial expenditure and opportunity costs (staff and volunteer time). HR costs include the time of government staff and community volunteers involved in planning, mobilisation, supervision, and delivery of MDA. Unit cost estimates were taken from Malawi costing studies (Appendix Table 4).

###### Linear scaling assumptions and their limitations

Applying full HR and programme unit costs proportionally to the number of people treated presumes a linear relationship between coverage and staffing requirements. This assumption is unlikely to hold in practice because: (i) many activities involve fixed or semi-fixed labour inputs; (ii) additional coverage can often be absorbed within existing staff capacity; (iii) economies of scope arise through shared delivery platforms; and (iv) supervisory and management structures do not expand in strict proportion to treatment numbers.

A fully proportional application of HR inputs therefore risks overstating the marginal cost of increased scale-up.

###### Three cost specifications for ICERs

To address these uncertainties and provide decision-makers with a transparent range of estimates, three costing specifications were used:

**Base case: Consumables plus financial costs for implementation (variable costs).**

Reflects the marginal cost of scaling up treatment

Excludes opportunity costs to avoid upward bias arising from linear scaling.

**Intermediate scenario: Consumables plus 50% of implementation costs.**

Applies half of the routine financial programme costs.

Represents a pragmatic midpoint assumption between fully variable and fully proportional scaling.

Intended to capture the plausible contribution of semi-fixed labour and delivery inputs without assuming full linear expansion.

**Minimal scenario: Consumables only.**

Represents the lower bound for costs, assuming no costs relating to delivery.

Included for completeness and sensitivity analysis, not as a reflection of marginal implementation costs.

The base-case ICERs provide the most conservative and policy-relevant estimate of the marginal cost of expanding MDA. The intermediate scenario illustrates how results change when a proportion of programme and staffing costs is incorporated, reflecting partial economies of scale. Consumables-only ICERs should be interpreted as lower-bound values, particularly relevant in settings where MDA platforms and staffing are already established.

Table 8. Maximum allowable costs (MAC) in USD per MDA treatment under alternative assumptions on WASH.

|  | **Pause WASH** | | | **Scale-up WASH** | | |
| --- | --- | --- | --- | --- | --- | --- |
| **District** | **SAC** | **PSAC+SAC** | **All** | **SAC** | **PSAC+SAC** | **All** |
| Balaka | 2.11 (1.54–2.67) | 0.78 (0.44–1.12) | 6.05 (4.41–7.68) | 1.28 (0.91–1.65) | 0.95 (0.67–1.23) | 3.69 (2.62–4.76) |
| Blantyre | 1.03 (0.59–1.46) | 0.40 (0.12–0.68) | 4.59 (2.70–6.48) | 0.55 (0.32–0.77) | 0.40 (0.23–0.56) | 2.49 (1.50–3.47) |
| Blantyre City | 0.55 (0.18–0.92) | 27.64 (26.51–28.77) | 2.51 (0.85–4.17) | 0.45 (0.27–0.63) | 0.32 (0.19–0.45) | 2.04 (1.25–2.83) |
| Chikwawa | 36.67 (35.09–38.24) | 17.40 (12.03–22.77) | 36.69 (34.78–38.60) | 33.97 (32.63–35.32) | 25.97 (25.06–26.88) | 34.18 (32.66–35.69) |
| Chiradzulu | 15.30 (10.54–20.07) | 0.67 (0.40–0.95) | 5.91 (4.03–7.78) | 7.40 (3.34–11.47) | 8.34 (3.81–12.88) | 2.80 (1.24–4.36) |
| Chitipa | 0.90 (0.54–1.26) | 22.39 (20.91–23.87) | 4.04 (2.47–5.62) | 0.44 (0.24–0.64) | 0.32 (0.17–0.47) | 2.04 (1.15–2.93) |
| Dedza | 29.83 (27.70–31.95) | 1.20 (0.91–1.48) | 34.63 (32.25–37.00) | 28.03 (26.03–30.04) | 21.50 (19.95–23.06) | 32.70 (30.36–35.04) |
| Dowa | 1.60 (1.23–1.98) | 1.23 (0.90–1.56) | 4.60 (3.48–5.71) | 1.21 (0.77–1.65) | 0.90 (0.57–1.22) | 3.50 (2.22–4.79) |
| Karonga | 1.64 (1.20–2.08) | 22.68 (20.97–24.38) | 4.75 (3.48–6.03) | 1.38 (0.88–1.89) | 1.03 (0.65–1.40) | 4.03 (2.59–5.46) |
| Kasungu | 29.90 (27.88–31.92) | 27.68 (25.98–29.38) | 34.77 (32.09–37.45) | 27.47 (25.54–29.40) | 20.87 (19.39–22.35) | 31.84 (29.52–34.16) |
| Likoma | 36.39 (34.26–38.52) | 1.27 (0.93–1.62) | 37.44 (35.01–39.87) | 32.82 (30.52–35.12) | 24.94 (23.20–26.68) | 33.82 (31.24–36.40) |
| Lilongwe | 1.69 (1.23–2.14) | 0.61 (0.30–0.92) | 4.88 (3.57–6.19) | 1.53 (1.01–2.05) | 1.14 (0.75–1.53) | 4.41 (2.92–5.91) |
| Lilongwe City | 0.83 (0.42–1.24) | 21.06 (19.77–22.35) | 2.42 (1.26–3.58) | 0.46 (0.23–0.69) | 0.33 (0.16–0.50) | 1.36 (0.71–2.01) |
| Machinga | 27.96 (26.28–29.65) | 26.61 (24.29–28.92) | 38.04 (35.54–40.54) | 25.32 (22.66–27.98) | 19.11 (17.10–21.13) | 34.55 (30.78–38.32) |
| Mangochi | 35.00 (32.15–37.85) | 1.30 (1.03–1.57) | 35.34 (32.35–38.33) | 31.17 (28.86–33.48) | 23.44 (21.68–25.20) | 31.32 (28.87–33.78) |
| Mchinji | 1.75 (1.39–2.10) | 17.22 (15.24–19.19) | 5.00 (3.98–6.03) | 1.38 (0.93–1.83) | 1.03 (0.69–1.36) | 3.97 (2.69–5.25) |
| Mulanje | 22.84 (20.25–25.44) | 0.57 (0.40–0.74) | 30.78 (27.07–34.48) | 20.09 (17.67–22.51) | 15.19 (13.27–17.12) | 27.12 (23.62–30.62) |
| Mwanza | 0.73 (0.52–0.95) | 0.67 (0.48–0.86) | 3.35 (2.39–4.31) | 0.53 (0.18–0.88) | 0.40 (0.12–0.67) | 2.45 (0.92–3.97) |
| Mzimba | 0.89 (0.64–1.15) | 0.37 (0.22–0.52) | 3.98 (2.90–5.05) | 0.72 (0.43–1.01) | 0.52 (0.31–0.74) | 3.19 (1.97–4.40) |
| Mzuzu City | 0.51 (0.31–0.71) | 1.60 (1.18–2.02) | 2.34 (1.46–3.21) | 0.27 (0.14–0.40) | 0.19 (0.09–0.30) | 1.31 (0.72–1.91) |
| Neno | 2.13 (1.57–2.70) | 1.33 (1.06–1.59) | 6.13 (4.46–7.79) | 1.57 (1.21–1.92) | 1.17 (0.90–1.45) | 4.51 (3.50–5.52) |
| Nkhata Bay | 1.77 (1.40–2.13) | 22.06 (19.45–24.67) | 5.05 (4.04–6.05) | 0.96 (0.69–1.23) | 0.72 (0.51–0.93) | 2.80 (2.02–3.57) |
| Nsanje | 29.24 (25.91–32.57) | 1.30 (0.98–1.61) | 33.63 (29.50–37.75) | 25.67 (22.95–28.39) | 19.37 (17.20–21.54) | 29.78 (26.36–33.21) |
| Ntcheu | 1.72 (1.31–2.13) | 0.76 (0.54–0.98) | 4.95 (3.77–6.14) | 1.60 (1.12–2.09) | 1.19 (0.83–1.54) | 4.62 (3.23–6.02) |
| Ntchisi | 1.03 (0.74–1.32) | 24.15 (22.95–25.36) | 3.00 (2.16–3.85) | 0.64 (0.29–1.00) | 0.48 (0.21–0.75) | 1.91 (0.87–2.94) |
| Phalombe | 31.70 (30.28–33.12) | 1.12 (0.83–1.41) | 37.09 (35.17–39.02) | 26.37 (24.71–28.02) | 20.06 (18.73–21.39) | 30.79 (28.66–32.91) |
| Rumphi | 1.50 (1.11–1.88) | 1.31 (0.84–1.78) | 4.34 (3.25–5.43) | 0.85 (0.53–1.18) | 0.63 (0.38–0.89) | 2.49 (1.56–3.42) |
| Thyolo | 1.52 (0.99–2.05) | 24.12 (22.61–25.62) | 4.34 (2.84–5.83) | 1.14 (0.56–1.72) | 0.94 (0.46–1.42) | 3.27 (1.64–4.90) |
| Zomba | 31.65 (29.70–33.60) | 18.87 (17.53–20.21) | 36.89 (34.48–39.31) | 28.30 (25.53–31.08) | 21.39 (19.18–23.60) | 32.79 (29.45–36.12) |
| Zomba City | 24.98 (23.20–26.75) | 0.78 (0.44–1.12) | 29.30 (27.18–31.41) | 24.24 (22.63–25.85) | 18.26 (17.11–19.41) | 28.23 (26.35–30.10) |

These values represent fiscal headroom for implementation beyond consumables at a threshold of USD 88 per DALY averted over 2024-2050.

Table 9. Disability-adjusted life years averted for each MDA strategy across alternative WASH assumptions.

| **Draw** | **mean** | **lower** | **upper** |
| --- | --- | --- | --- |
| Continue WASH | | | |
| MDA All | 18,140,689.83 | 17,758,818.61 | 18,522,561.06 |
| MDA PSAC+SAC | 18,042,676.52 | 17,658,548.78 | 18,426,804.26 |
| MDA SAC | 17,998,129.72 | 17,614,644.15 | 18,381,615.28 |
| Pause WASH | | | |
| MDA All | 18,108,731.73 | 17,764,994.54 | 18,452,468.92 |
| MDA PSAC+SAC | 18,011,564.35 | 17,664,035.69 | 18,359,093.00 |
| MDA SAC | 17,968,093.95 | 17,622,264.05 | 18,313,923.85 |
| Scale-up WASH | | | |
| MDA All | 16,151,421.34 | 15,711,079.35 | 16,591,763.34 |
| MDA PSAC+SAC | 16,053,916.53 | 15,613,512.12 | 16,494,320.93 |
| MDA SAC | 16,009,295.73 | 15,568,518.38 | 16,450,073.09 |

Values are weighted by district and summed across 2024-2050. Lower and upper denote the 95% confidence interval bounds.

Table 10. Number and Proportion of Districts by Cost-Effectiveness Category for alternative MDA strategies across sensitivity analyses.

|  | Comparator | Cost-saving | Cost-effective | Not cost-effective | Dominated |
| --- | --- | --- | --- | --- | --- |
| Main analysis | | | | | |
|  | SAC vs no MDA | 0 (0.0%) | 29 (90.6%) | 3 (9.4%) | 0 (0.0%) |
|  | PSAC+SAC vs SAC | 0 (0.0%) | 1 (3.1%) | 9 (28.1%) | 22 (68.8%) |
|  | All vs PSAC+SAC | 11 (34.4%) | 0 (0.0%) | 0 (0.0%) | 21 (65.6%) |
| Alternative WASH assumptions | | | | | |
| Pause WASH | SAC vs no MDA | 0 (0.0%) | 30 (93.8%) | 2 (6.2%) | 0 (0.0%) |
|  | PSAC+SAC vs SAC | 0 (0.0%) | 1 (3.1%) | 10 (31.2%) | 21 (65.6%) |
|  | All vs PSAC+SAC | 11 (34.4%) | 0 (0.0%) | 0 (0.0%) | 21 (65.6%) |
| Scale-up WASH | SAC vs no MDA | 10 (31.2%) | 0 (0.0%) | 0 (0.0%) | 22 (68.8%) |
|  | PSAC+SAC vs SAC | 0 (0.0%) | 1 (3.1%) | 9 (28.1%) | 22 (68.8%) |
|  | All vs PSAC+SAC | 0 (0.0%) | 25 (78.1%) | 7 (21.9%) | 0 (0.0%) |
| Alternative discounting assumptions | | | | | |
| 3% annual discount for costs | SAC vs no MDA | 0 (0.0%) | 31 (96.9%) | 1 (3.1%) | 0 (0.0%) |
|  | PSAC+SAC vs SAC | 0 (0.0%) | 2 (6.2%) | 8 (25.0%) | 22 (68.8%) |
|  | All vs PSAC+SAC | 10 (31.2%) | 1 (3.1%) | 0 (0.0%) | 21 (65.6%) |
| 3% annual discount for health | SAC vs no MDA | 0 (0.0%) | 25 (78.1%) | 7 (21.9%) | 0 (0.0%) |
|  | PSAC+SAC vs SAC | 0 (0.0%) | 1 (3.1%) | 9 (28.1%) | 22 (68.8%) |
|  | All vs PSAC+SAC | 11 (34.4%) | 0 (0.0%) | 0 (0.0%) | 21 (65.6%) |
| Alternative costing assumptions | | | | | |
| Consumables plus 0.5 * implementation costs | SAC vs no MDA | 0 (0.0%) | 31 (96.9%) | 1 (3.1%) | 0 (0.0%) |
|  | PSAC+SAC vs SAC | 0 (0.0%) | 3 (9.4%) | 7 (21.9%) | 22 (68.8%) |
|  | All vs PSAC+SAC | 11 (34.4%) | 0 (0.0%) | 0 (0.0%) | 21 (65.6%) |
| Consumables only | SAC vs no MDA | 0 (0.0%) | 32 (100.0%) | 0 (0.0%) | 0 (0.0%) |
|  | PSAC+SAC vs SAC | 0 (0.0%) | 10 (31.2%) | 0 (0.0%) | 22 (68.8%) |
|  | All vs PSAC+SAC | 2 (6.2%) | 9 (28.1%) | 0 (0.0%) | 21 (65.6%) |
| Alternative disability weight assumptions | | | | | |
| Disability weights assigned to light infections | SAC vs no MDA | 0 (0.0%) | 31 (96.9%) | 1 (3.1%) | 0 (0.0%) |
|  | PSAC+SAC vs SAC | 2 (6.2%) | 3 (9.4%) | 16 (50.0%) | 11 (34.4%) |
|  | All vs PSAC+SAC | 29 (90.6%) | 2 (6.2%) | 1 (3.1%) | 0 (0.0%) |

*National-level results are summarised as the number and percentage of districts classified into cost-effectiveness decision categories for each intervention comparison and scenario. Decision categories were defined using incremental cost-effectiveness analysis relative to the specified comparator as follows: Cost-saving indicates the intervention generated greater health benefit at lower cost (dominant); Cost-effective (≤88 USD) indicates the intervention produced additional health benefit at higher cost with an incremental cost-effectiveness ratio (ICER) below the cost-effectiveness threshold; Not cost-effective indicates additional health benefit at higher cost with an ICER exceeding the threshold; and Dominated indicates the intervention was either more costly and less effective or produced no additional health benefit relative to the comparator.*

*Table 9. DALYs incurred under different assumptions on DALY weights for light infection*

|  | **No MDA** | **MDA SAC** | **MDA PSAC+SAC** | **MDA All** | **No MDA** |
| --- | --- | --- | --- | --- | --- |
| Light infection disability included | 19.28  (18.53–20.38) | 0.73  (0.68–0.79) | 0.64  (0.60–0.69) | 0.06  (0.05–0.06) | 17.28  (16.36–18.26) |
| Light infection disability excluded *(Main analysis)* | 18.17  (17.42–19.29) | 0.18  (0.14–0.22) | 0.13  (0.10–0.17) | 0.03  (0.03–0.04) | 16.19  (15.26–17.17) |

*Values are in millions of DALYs incurred over the simulation period 2024-2050, weighted by district.*

##### Optimal MDA strategy as a function of the opportunity cost threshold

We quantified how the net health benefit (NHB)-maximising MDA strategy varies with the opportunity cost threshold λ (USD per DALY). For each λ ∈ (20, 40, 60, 80, … 600)we computed, for every district and strategy under the Continue WASH assumption,

$${NHB}_{\lambda}=\lambda\cdot DALYs averted-Cost$$

using outcomes versus no MDA, aggregated over 2024-2050 without discounting. For each district, the strategy with the highest mean NHB was selected, then the proportion of districts for which each strategy was optimal was determined.

Figure 1 illustrates how the proportion of Malawi districts for which each MDA strategy is optimal varies as the willingness-to-pay threshold (λ) increases. For each value of λ, net health benefit was calculated for all strategies relative to no MDA, and the strategy with the highest value was identified for every district. The three curves therefore depict the share of districts in which annual SAC-only MDA, annual PSAC + SAC MDA, or annual all-age MDA yields the greatest net health benefit at each λ.

When incorporating the programme financial costs, the MDA All strategy was NHB-optimal in the majority of districts across all willingness-to-pay thresholds examined. Even at relatively low thresholds, MDA All maximised NHB in approximately 80-90% of districts, with only a small proportion favouring less expansive strategies. MDA SAC was optimal in a minority of districts at lower thresholds, reflecting settings where the incremental costs of expanding treatment coverage outweighed additional health gains under constrained willingness-to-pay assumptions. The intermediate strategy (MDA PSAC+SAC) was rarely optimal, indicating limited settings in which partial expansion represented the efficiency-maximising option.

When only consumables costs were considered (Figure 1b), the relative distribution of optimal strategies shifted markedly. Under this costing assumption, MDA SAC was optimal in a substantially larger proportion of districts at lower opportunity cost thresholds, accounting for approximately 30-40% of districts near the lower threshold. This pattern reflects the increased relative weight of incremental treatment costs once fixed or shared programme implementation costs are removed, thereby reducing the marginal efficiency of expanding coverage under stringent cost-effectiveness thresholds. Nonetheless, as λ increased, the proportion of districts favouring expanded strategies rose, with MDA All again becoming the dominant strategy across most districts at higher thresholds.

Across both costing assumptions, the intermediate expansion strategy (MDA PSAC+SAC) remained infrequently optimal, suggesting that incremental expansion to preschool aged children alone may offer limited efficiency advantages compared with either maintaining school aged treatment or expanding directly to universal coverage over this timeframe (2024-2050). The choice of costing perspective substantially influences the estimated optimal policy under constrained willingness-to-pay thresholds, while the relative robustness of the MDA All strategy across thresholds suggests strong economic justification for expanded treatment in most settings.

*(a)*

*(b)*
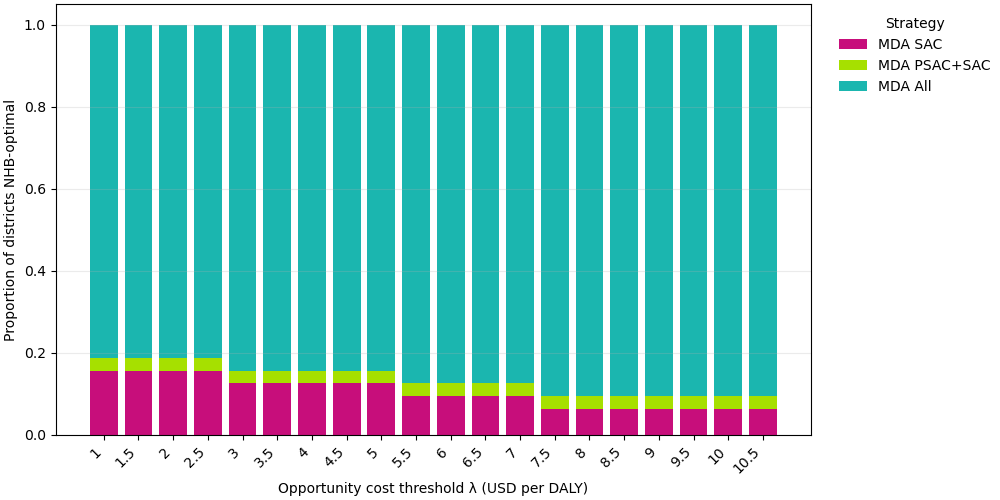

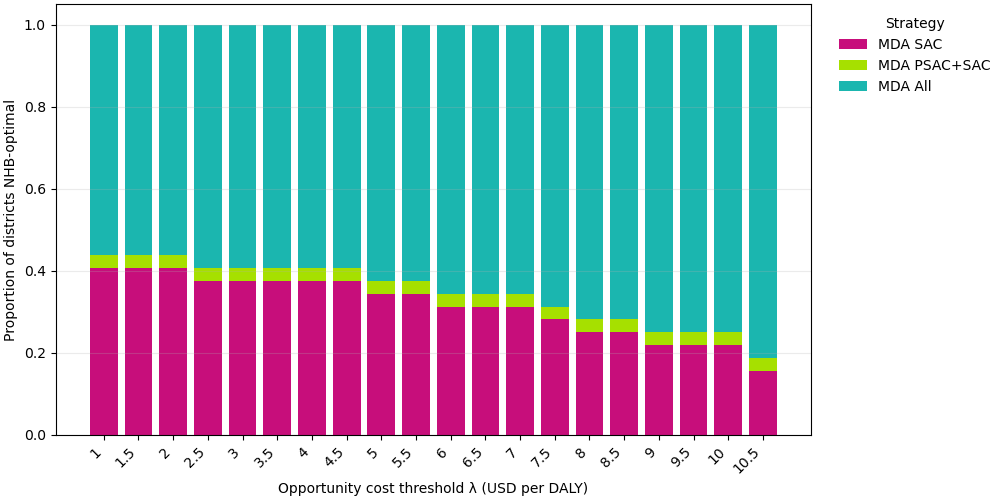

Figure 3. Optimal MDA strategy by willingness-to-pay threshold (λ): proportion of districts in which each strategy maximises net health benefit.

Financial costs (implementation plus consumables) are included in (a) while (b) uses only the consumables costs required for MDA programmes.
